# Antigenic responses are hallmarks of fibrotic interstitial lung diseases independent of underlying etiologies

**DOI:** 10.1101/2023.05.08.23289640

**Authors:** Young me Yoon, Tania E. Velez, Vaibhav Upadhyay, Sara E. Vazquez, Cathryn T. Lee, Kavitha C. Selvan, Christopher S. Law, Kelly M. Blaine, Maile K. Hollinger, Donna C. Decker, Marcus R. Clark, Mary E. Strek, Robert D. Guzy, Ayodeji Adegunsoye, Imre Noth, Paul J. Wolters, Mark Anderson, Joseph L. DeRisi, Anthony K. Shum, Anne I. Sperling

**Affiliations:** University of Chicago, Department of Medicine, Chicago, IL 60637; University of Virginia, Department of Medicine, Charlottesville, VA 22908; University of California San Francisco, Department of Medicine, San Francisco, CA 94143; University of California San Francisco and Chan Zuckerberg Biohub, San Francisco, CA 94158; University of Wisconsin at Madison, Department of Medicine, Madison, WI 53792

## Abstract

Interstitial lung diseases (ILD) are heterogeneous conditions that may lead to progressive fibrosis and death of affected individuals. Despite diversity in clinical manifestations, enlargement of lung-associated lymph nodes (LLN) in fibrotic ILD patients predicts worse survival. Herein, we revealed a common adaptive immune landscape in LLNs of all ILD patients, characterized by highly activated germinal centers and antigen-activated T cells including regulatory T cells (Tregs). In support of these findings, we identified serum reactivity to 17 candidate auto-antigens in ILD patients through a proteome-wide screening using phage immunoprecipitation sequencing. Autoantibody responses to actin binding LIM protein 1 (ABLIM1), a protein highly expressed in aberrant basaloid cells of fibrotic lungs, were correlated with LLN frequencies of T follicular helper cells and Tregs in ILD patients. Together, we demonstrate that end-stage ILD patients have converging immune mechanisms, in part driven by antigen-specific immune responses, which may contribute to disease progression.

## Introduction

Interstitial lung disease (ILD) is a heterogeneous group of conditions characterized by inflammation and scarring of the lung parenchyma. Diagnosis and treatment strategies for ILD subtypes have been largely based on the underlying etiology. The most common type of ILD is idiopathic pulmonary fibrosis (IPF). IPF is diagnosed based on the presence of a specific radiologic or histopathologic pattern, usual interstitial pneumonia, in the absence of an identifiable underlying cause (Raghu et al., 2022). In addition to IPF, several other ILDs including hypersensitivity pneumonitis (HP) and connective tissue disease associated ILD (CTD-ILD) can also progress to pulmonary fibrosis (Kern et al., 2015; Yusen et al., 2014). HP is driven by inflammatory responses against inhaled environmental antigens, such as molds and bird droppings^9^. Other pulmonary fibrosis patients have either underlying CTD-ILD^10^ or features of autoimmunity, including serum autoantibodies, without meeting the full diagnostic criteria for a connective tissue disease (Ghang et al., 2019; Oldham et al., 2016). Immunosuppressants are commonly used to improve symptoms and slow disease progression in patients with HP and other immune-associated ILD. In contrast, broadly suppressing the immune system in IPF patients has been ineffective at improving patient outcomes, and instead increased the risk for hospitalization and death (Idiopathic Pulmonary Fibrosis Clinical Research et al., 2012). As a result, the prevailing mechanistic paradigm is that IPF is driven by epithelial dysfunction, while HP and CTD-ILD are proposed to be driven by immune mechanisms (Wolters et al., 2018).

While the immune system may not be the primary etiologic driver of IPF, there is growing evidence of abnormal immune activation in fibrotic ILDs, including IPF. For example, many immune-associated receptors and ligands have been identified as biomarkers for IPF (DePianto et al., 2015; Herazo-Maya et al., 2013; Parra et al., 2007; Vuga et al., 2014), and activated T cells and B cells in IPF patient lungs negatively correlate with patient survival (Marchal-Somme et al., 2006; Todd et al., 2013). Furthermore, enlargement of lung-draining lymph nodes (LLN) is prevalent and predicts worse survival in patients with all types of ILDs, including IPF (Adegunsoye et al., 2019). As lymph nodes are structures where adaptive immune cells are recruited and proliferate in response to antigens from the tissues, the association between LLN size and patient survival suggest that the adaptive immune response could contribute to the pathophysiology of IPF. This idea is supported by findings that compared to healthy controls, IPF patients have higher levels of circulating autoantibodies (Feghali-Bostwick et al., 2007; Heukels et al., 2019). While efforts at characterizing patients with autoantibodies and no defined CTD-ILD are ongoing (Fischer et al., 2015), the extent of autoantibody specificities and their functional implications in fibrotic ILD are incompletely understood. Thus, determining specific immune pathways that impact disease progression and antigenic specificity of the response are critical for developing mechanistic understanding of pathology and effective therapeutic strategies for IPF and other fibrotic ILDs.

To understand the complexity of immune responses in fibrotic ILD, we investigated the phenotypes of adaptive immune cells in LLNs isolated from explanted lungs of end-stage ILD patients and age-matched controls. Strong immune activation profiles that are consistent with antigen-stimulation of T cells and germinal centers (GC) were identified across all ILD LLN, including IPF. Furthermore, Tregs were elevated and had unique phenotypes in ILD LLNs compared to controls, suggesting that failure to control antigen responses in ILD is potentially due to Treg dysregulation. To identify antigens that drive immune activation in ILD, we employed a human peptidome phage library to screen patients’ serum antibodies against autoantigens. We discovered 17 candidate autoantigens that are shared among ILD patients regardless of subtypes. Strikingly, many ILD patients have circulating antibodies against ABLIM1, an actin-binding protein that is specifically upregulated in structural cells of the fibrotic lungs, suggesting a common antigenic stimulation in fibrotic ILD patients. Thus, we demonstrate that all ILD patients’ enlarged lymph nodes contained evidence of antigen-induced activation that was dysregulated, and we identified a novel autoantigen that could be a part of common immunopathology in pulmonary fibrosis.

## Results

### Distinct T cell and B cell landscapes in ILD lung lymph nodes compared to age-matched controls

To investigate the enlarged lung lymph nodes in ILD patients, we conducted comprehensive profiling of adaptive immune cells in biobanked LLN from end-stage fibrotic ILD patients and organ donors who were free of known lung disease. For this study, groups were matched for age (median ages of 63 and 60 for ILD and controls, respectively) and sex (72% and 67% male) (Table 1). ILD subtype was determined via multidisciplinary diagnosis using ATS/ERS criteria (Fischer et al., 2015; Raghu et al., 2018; Raghu et al., 2020). IPF (n = 30) was most prevalent in our study cohort, followed by interstitial pneumonia with autoimmune features (IPAF: n = 7), CTD-ILD: (n = 5), HP: (n = 4), and other ILDs (n = 4). In the CTD-ILD group, 3 patients had rheumatoid arthritis (RA), 1 patient had anti-synthetase syndrome, and 1 patient had anti-neutrophil cytoplasmic antibody-associated ILD. The other ILD group included patients with short telomere syndrome, pleuroparenchymal fibroelastosis, and unclassifiable ILD.

**Table 1.** Demographics of explanted LLN flow cytometry study cohorts

|  |  | ILD (n=50) | Control (n=36) |
| --- | --- | --- | --- |
| **Sex, n (%)** | Male | 36 (72%) | 24 (67%) |
|  | Female | 14 (28%) | 12 (33%) |
| **Age, median (range)** |  | 63 (43 – 72) | 60 (44 – 76) |
| **Race, n (%)** | African American | 6 (12%) | 8 (22%) |
|  | Asian | 2 (4%) |  |
|  | Caucasian | 36 (72%) | 26 (72%) |
|  | Hispanic | 1 (2%) | 2 (6%) |
|  | Other | 1 (2%) |  |
|  | Unknown | 4 (8%) |  |
| **Diagnosis, n (%)** | IPF | 30 (60%) |  |
|  | IPAF | 7 (14%) |  |
|  | CTD-ILD | 5 (10%) |  |
|  | HP | 4 (8%) |  |
|  | Other ILD<sup>1</sup> | 4 (8%) |  |

<sup>1</sup>Other ILD included patients with short telomere syndrome, pleuroparenchymal fibroelastosis, and unclassifiable ILD.

We designed two antibody panels for spectral flow cytometry to investigate compositions and phenotypes of T cells and B cells. Due to the large number of samples, flow cytometry was performed in 4 batches on different days. Batches 1-3 consisted of ILD samples with age matched controls, and batch 4 consisted of ILD samples only (IPF and HP). We found that ILD patients and control donors had similar levels of total T cells in their LLN (Fig. 1 A and B). No significant difference was found based on different types of ILDs. However, principal component analysis (PCA) using 19 T cell markers revealed a clear separation between ILD and control groups, consistently across batches (Fig. 1 C and E). To investigate which markers are associated with the distinct T cell phenotypes, we performed logistic regression analyses across all samples, with adjustments for batch effects, and found that expression of 11 markers were significantly different between ILD and control groups (Fig. 1 F; Fig. S1 A). Interestingly, costimulatory molecule CD154 (CD40L) and regulatory T cell (Treg) marker FOXP3 were among the most significantly associated and highly expressed markers in ILD patient T cells. Other markers associated with activation (HLA-DR and ICOS) and Th2-lineage determining transcription factor GATA3 were also upregulated in ILD T cells. In contrast, significantly lower CCR7 expression in ILD LLN T cells suggested that these T cells are poised to exit lymph nodes and traffic to the lungs. Expressions of cytokine receptors CD25 (IL-2R) and CD127 (IL-7R) were also downregulated in ILD T cells. To visualize protein expression patterns at the cellular level, we performed a uniform manifold approximation and projection (UMAP) with T cells from batch 1 (Fig. S1, B and C). We found that CD154 and FOXP3 were not co-expressed, while FOXP3-high cells also expressed CD25, ICOS, HLA-DR, and CD137 (Fig. S1 C). Furthermore, some ICOS-positive cells co-expressed PD1 and BCL6, which are consistent with the follicular helper T cell (Tfh) phenotype. Overall, these data suggest that T cells in ILD LLNs have unique phenotypes compared to controls.

**Figure 1.**
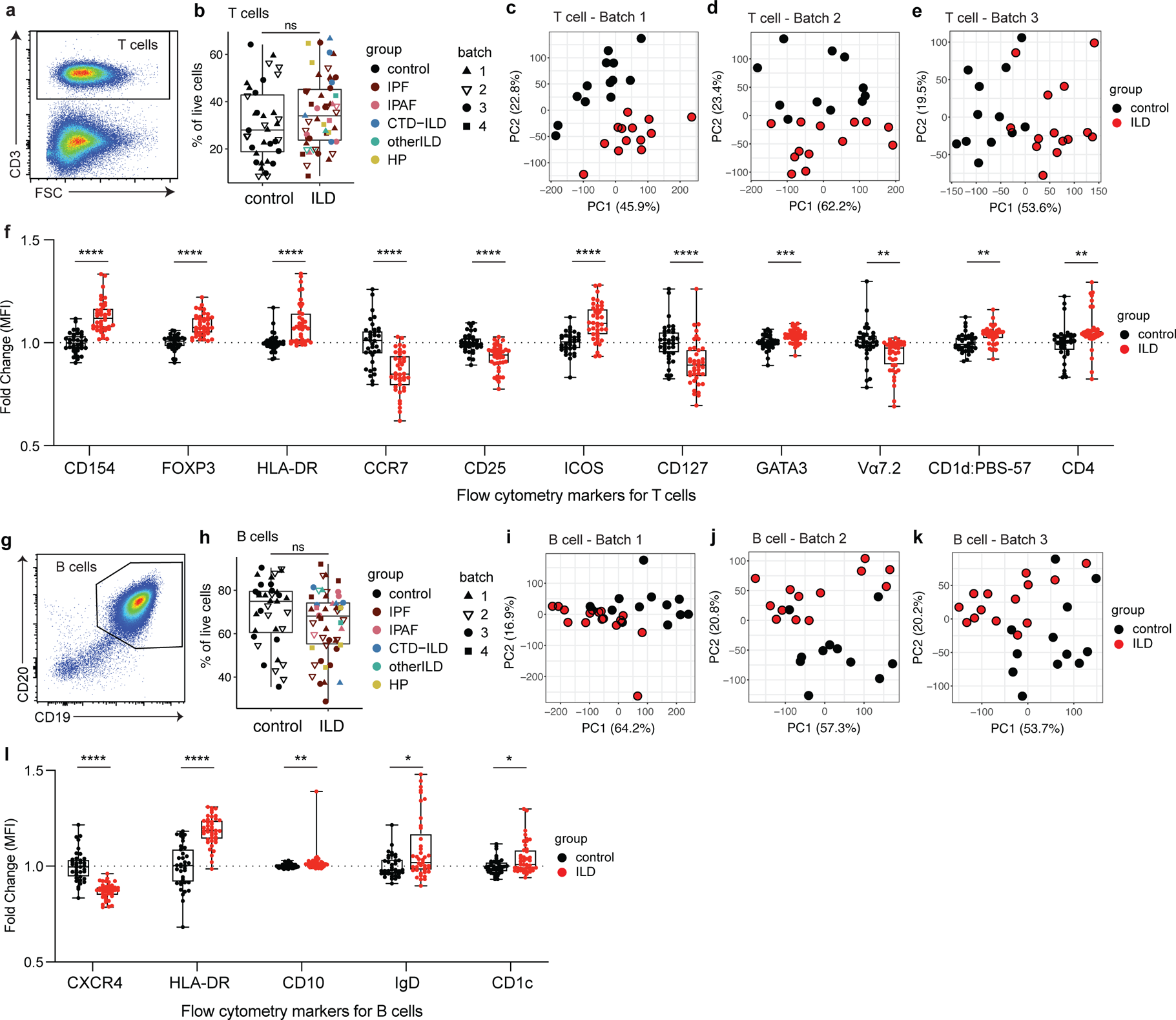
Distinct T cell and B cell landscapes in LLNs from ILD patients compared with matched controls. **a.** Representative flow cytometry gating for total T cells in LLNs. Cells were pre-gated on live cells. **b.** Frequencies of T cells in live cells in control vs ILD LLNs. Statistical significance was tested by the Wilcoxon rank sum test. Differential diagnosis and experimental batch are noted by the color and symbol as shown in the legend. **c-e.** PCA of total T cells based on fluorescence intensities of 19 parameters: CD154, FOXP3, HLA-DR, CCR7, CD25, ICOS, CD127, GATA3, Vα7.2, PBS-57 loaded CD1d tetramer, CD4, CD8, Vδ, PD1, RORγt, BCL6, CD38, CD45RA and CD137. An analysis was run with 10,000 cells, 8,000 cells, and 10,000 cells for batch1, batch2, and batch 3, respectively. **f.** Box plots comparing the median fluorescence intensity of individual T cell markers between ILD and controls, based on all samples in batches 1-3. Fold change was calculated relative to the mean values of control samples, separately for each batch. Parameters that are significantly associated with ILD based on a likelihood-ratio test on each logistic regression coefficient are shown. Non-significant parameters are shown in Extended Figure 1. P-values were adjusted by Benjamini-Hochberg correction. **g.** Representative flow cytometry gating for B cells in LLNs. Cells were pre-gated on live cells. **h.** Frequencies of B cells in live cells in control vs ILD LLNs. Statistical significance was tested by the Wilcoxon rank sum test. **i-k**. PCA of B cells based on fluorescence intensities of 12 parameters: CXCR4, HLA-DR, CD10, IgD, CD1c, CD44, CD138, CD11C, CD24, CD27, CD38, IgM. An analysis was run with 10,000 cells for each batch, separately. **l.** Box plots comparing the median fluorescence intensity of individual B cell markers between ILD and controls, based on all samples in batches 1-3. Calculations of fold changes and adjusted p-values were done as described above (f). P<0.05 (*), P<0.01 (**), P<0.001 (***), P<0.0001 (****).

Similar to T cells, there was no significant difference in the frequency of total B cells in the LLN between ILD and controls (Fig. 1, G and H). PCA analysis of B cells based on 12 parameters demonstrated that distinct B cell phenotypes exist in ILD LLNs compared to controls (Fig. 1, I and K). Based on regression models, we found 5 B cell markers are significantly associated with ILD (Fig. 1 l; and Fig. S1 D). Expression of CXCR4 was the most significant association and was the only marker downregulated in ILD patients. Other markers upregulated in ILD patients included HLA-DR, CD10, IgD, and CD1c. UMAP analysis of the B cells revealed differential islands of cells between the two groups (Fig S1, E and F). CD10 expression was confined to one region where there was high expression for CD38, which is known to be highly expressed on GC B cells and plasma cells in LN (Fig. S1 F). These data indicate that B cells also have distinct phenotypes in ILD patients compared to controls.

### Effector T cells and Tregs display activated phenotypes in ILD LLN

To interrogate which T cell subsets are driving the ILD phenotype, we analyzed CD4 and CD8 T cells separately. Frequencies of total CD4 T cells were trending up in ILD patients (Fig. S2 A), and CD8 T cells were trending down (Fig. S2 B), leading to a significantly higher ratio of CD4 T cells to CD8 T cells (Fig. S2C). The expression of several activation markers including HLA-DR on CD8 T cells were associated with ILD (Fig. S2, D and H). For CD4 T cells, we analyzed CD4 T conventional cells (Tcon) (Fig. 2, A-F) and Tregs (Fig. 2, G-I) separately since these two populations play opposite roles in immune response. Interestingly, PCA of CD4 Tcon cell phenotypes distinguished ILD patients from controls (Fig. 2, C-E), and ILD patients showed higher expression of activation markers CD154, HLA-DR, and ICOS on Tcon cells (Fig. 2 F; and Fig. S2 I). Unlike Tcon and CD8 T cell populations in LLN, Treg proportions were elevated in ILD LLNs compared to controls (Fig. 2, G and H). As Tregs can express a variety of effector transcription factors and activation markers that identify their function, we performed PCA and found that Tregs from the LLN of ILD patients have phenotypes that are distinct from control LLNs (Fig. 2, I-K). Intriguingly, expression of 7 markers were associated with ILD, five of which are activation markers such as PD1, HLA-DR, CD137, ICOS and CD154 (Fig. 2 l). Surprisingly, CD25 was the most significantly associated with ILD and was downregulated in ILD patients’ LLN compared to controls. Similar to total CD4 T cells, Tregs from ILD patients had reduced expression of CCR7, indicating that these cells were poised to leave the LLN and migrate back to the tissue (Fig. 2 l). These findings indicate that effector T cells and Tregs accumulate in ILD LLNs at end-stage disease, have distinct phenotypes, and therefore may have different functional roles compared to non-diseased organ donors.

**Figure 2.**
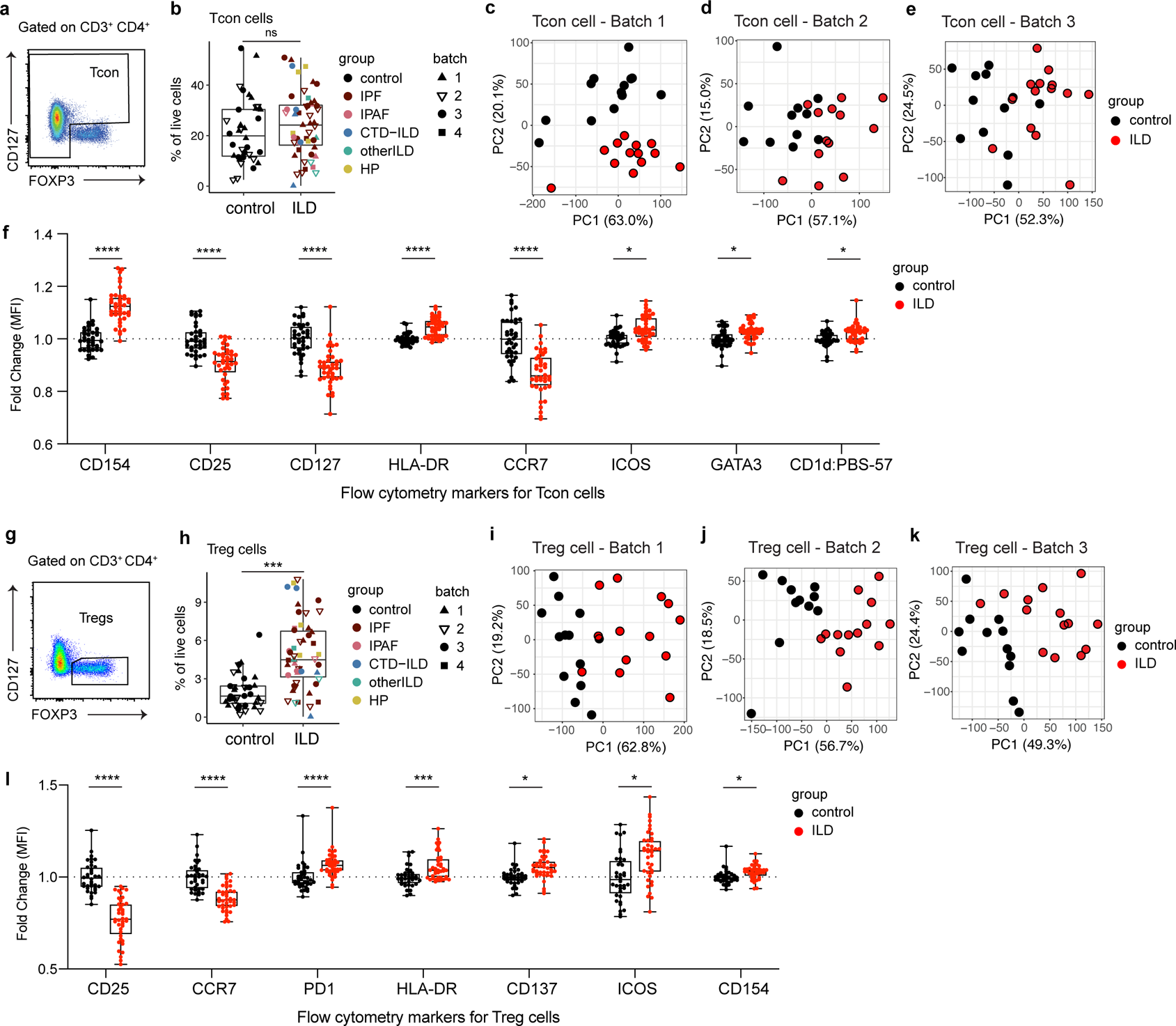
ILD patients have elevated Tregs with unique phenotypes compared to controls. **a.** Representative flow cytometry gating for Tcon cells in LLNs. Cells were pre-gated on live, CD3+, CD4+ cells. **b.** Frequencies of Tcon cells in live cells in control vs ILD LLNs. Statistical significance was tested by the Wilcoxon rank sum test. **c-e.** PCA of Tcon cells based on fluorescence intensities of 16 parameters: CD154, CD25, CD127, HLA-DR, CCR7, ICOS, GATA3, PBS-57 loaded CD1d tetramer, Vα7.2, RORγt, BCL6, PD1, CD137, CD38, CD45RA, and Vδ. An analysis was run with 10,000 cells, 2,300 cells, and 10,000 cells for batch1, batch2, and batch 3, respectively. **f.** Box plots comparing the median fluorescence intensity of individual Tcon markers between ILD and controls, based on all samples in batches 1-3. Fold change was calculated relative to the mean values of control samples, separately for each batch. Parameters that are significantly associated with ILD based on a likelihood-ratio test on each logistic regression coefficient are shown. Non-significant parameters are shown in Extended Figure 2. P-values were adjusted by Benjamini-Hochberg correction. **G.** Representative flow cytometry gating for Treg cells in LLNs. Cells were pre-gated on live, CD3+, CD4+ cells. **H.** Frequencies of Treg cells in live cells in control vs ILD LLNs. Statistical significance was tested by the Wilcoxon rank sum test. **I-k.** PCA of Treg cells based on fluorescence intensities of 12 parameters: CD25, CD154, HLA-DR, CCR7, ICOS, GATA3, RORγt, BCL6, PD1, CD137, CD38, CD45RA. An analysis was run with 1,000 cells, 269 cells, and 1,300 cells for batch1, batch2, and batch 3, respectively. **L.** Box plots comparing the median fluorescence intensity of individual Treg markers between ILD and controls, based on all samples in batches 1-3. Calculations of fold changes and adjusted p-values were done as described above (f). P<0.05 (*), P<0.01 (**), P<0.001 (***), P<0.0001 (****).

### Adaptive immune activation signatures define the ILD lymph node landscape

Using conventional flow cytometry gating, we identified and quantified a total of 39 unique T cell and B cell populations in LLNs (Fig. S3; and Fig. S4). Hierarchical clustering based on frequencies of individual immune cell populations demonstrated that almost all ILD samples clustered away from controls, suggesting specific immune cells are correlated with ILD (Fig. S5). Comparisons of cellular abundance between ILD and control LLNs determined that 25 cell populations were differentially enriched between the two groups (Fig. 3 A). Immune populations expanded in ILD LLNs were Tcon cells expressing co-stimulatory molecules CD154, ICOS, and PD1. Consistent with immune activation, T cells with effector memory phenotypes (EM of Tcon, TEMRA of Tcon, EM of CD8) were more abundant in ILD LLNs (Fig. 3 A). Furthermore, robust enrichment of Tfh cells and GC B cells indicate activated germinal center response in ILD LLN. In addition, total Tregs and a CD25-low subset of Tregs were highly enriched in ILD LLN. Only 4 populations were downregulated in ILD LLNs: central memory CD4 T cells, naïve CD8 T cells, switched memory B cells and Th2 cells. These data suggest that expansion of effector memory T cell proportions occurred at the expense of central memory and naïve T cells, which could support a hypothesis that there are ongoing immune responses in the lymph nodes of ILD patients.

**Figure 3.**
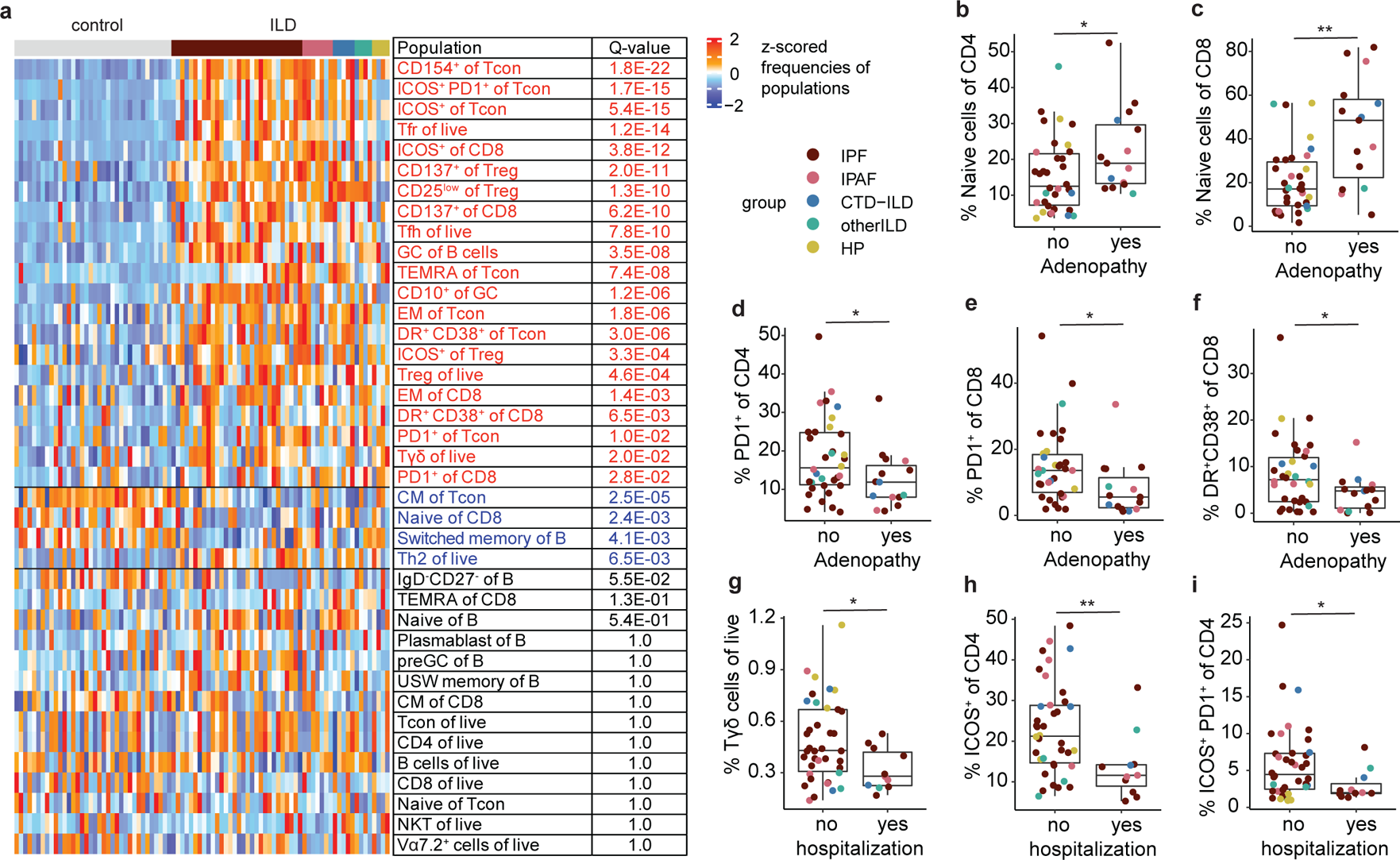
Cells in LLNs of ILD patients have significant adaptive immune activation signatures, some of which are associated with patients’ clinical history. **a.** Heatmap visualizing batch-normalized z-scores of populational frequencies ranked in the order of statistical significance in difference between ILD and control LLNs. A total of 39 populations were quantified by flow cytometry using gating strategies shown in Extended Figures 3 and 4. An associated with ILD was tested for each cell population by a likelihood-ratio test on each linear regression coefficient. Q-values are adjusted p-values based on the Benjamini-Hochberg method. **b-f.** Quantifications of immune cell populations that are significantly different between ILD patients who had lymphadenopathy (yes) and ILD patients who did not have lymphadenopathy (no) based on chest CT scans obtained at diagnosis. **g-i.** Quantifications of immune cell populations that are significantly different between ILD patients who had history of hospitalization due to respiratory cause (yes) and ILD patients who had not been hospitalized prior to transplantation (no).

### T cell phenotypes are associated with clinical phenotypes

Given that enlargement of LLNs at diagnosis correlates with survival (Adegunsoye et al., 2019), we next investigated whether the presence of lymphadenopathy at the time of diagnosis associates with accumulation of immune cell types in LLNs, obtained from each patient at the time of lung transplantation. Surprisingly, we found that naïve T cells, both CD4 and CD8 cells, are more abundant in LLNs of ILD patients who presented at diagnosis with lymphadenopathy compared to patients who did not have enlarged lymph nodes (Fig. 3, B and C). Increase in naïve T cells in enlarged LLN were accompanied by a decrease in PD1 expression on T cells, which is a marker of activation and exhaustion (McLane et al., 2019) (Fig. 3, D and E). Furthermore, another activation phenotype, manifest as the co-expression of HLA-DR and CD38, was lower in ILD patients who presented with lymphadenopathy compared to patients who did not have enlarged lymph nodes (Fig. 3 F). Thus, our data show that patients who had LLN enlargement at the time of diagnosis accumulate more naïve T cells and less PD-1 expressing T cells in their LLNs by the end stage of the disease.

As the frequency and etiology of hospitalizations are linked with survival in fibrotic ILD (Kim et al., 2021; Salonen et al., 2020), we tested whether ILD patients with history of hospitalization due to respiratory causes have different LLN immune profiles than other ILD patients. ILD patients with at least one hospitalization had significantly decreased Tγδ cells and ICOS^+^ CD4 T cells compared to ILD patients who had never been hospitalized (Fig. 3, G-I). These data suggest that recruitment of Tγδ cells and ICOS^+^ CD4 T cells may be protective for respiratory health.

### IPF and immune-associated ILD patients’ LLN T cells are similarly activated and expanded

The etiology of HP is known to be due to environmental exposures that induce immune responses, and CTD-ILD and IPAF are caused by, or associated with, autoimmune mechanisms. Therefore, involvement of the LLNs in immune-associated ILDs (immune-ILD) is not surprising. However, whether IPF specifically is associated with a dysregulated immune response, and whether the immune system plays any role in IPF etiology or pathology remains controversial. To test the hypothesis that IPF patients’ LLNs have different immune phenotypes compared to immune-ILD patients, we compared different T cell population frequencies between IPF and immune-ILD groups, as well as donor controls. Surprisingly, CD4 T cells in both IPF and immune-ILD LLNs significantly upregulated CD154 compared control LLNs (Fig. 4, A and B). Similarly, CD137 was upregulated on CD8 T cells (Fig. 4, C and D) and Tregs (Fig. 4, E and F) of both ILD groups compared to non-diseased control LLN. CD154 and CD137 are known to be upregulated in response to recent antigen stimulation of T cell receptors (Frentsch et al., 2005; Wolfl et al., 2007). Thus, both CD4 and CD8 T cells in the IPF LLNs were activated similarly to the immune-ILD group, suggesting that T cell activation may be down-stream of antigen-specific responses in IPF.

**Figure 4.**
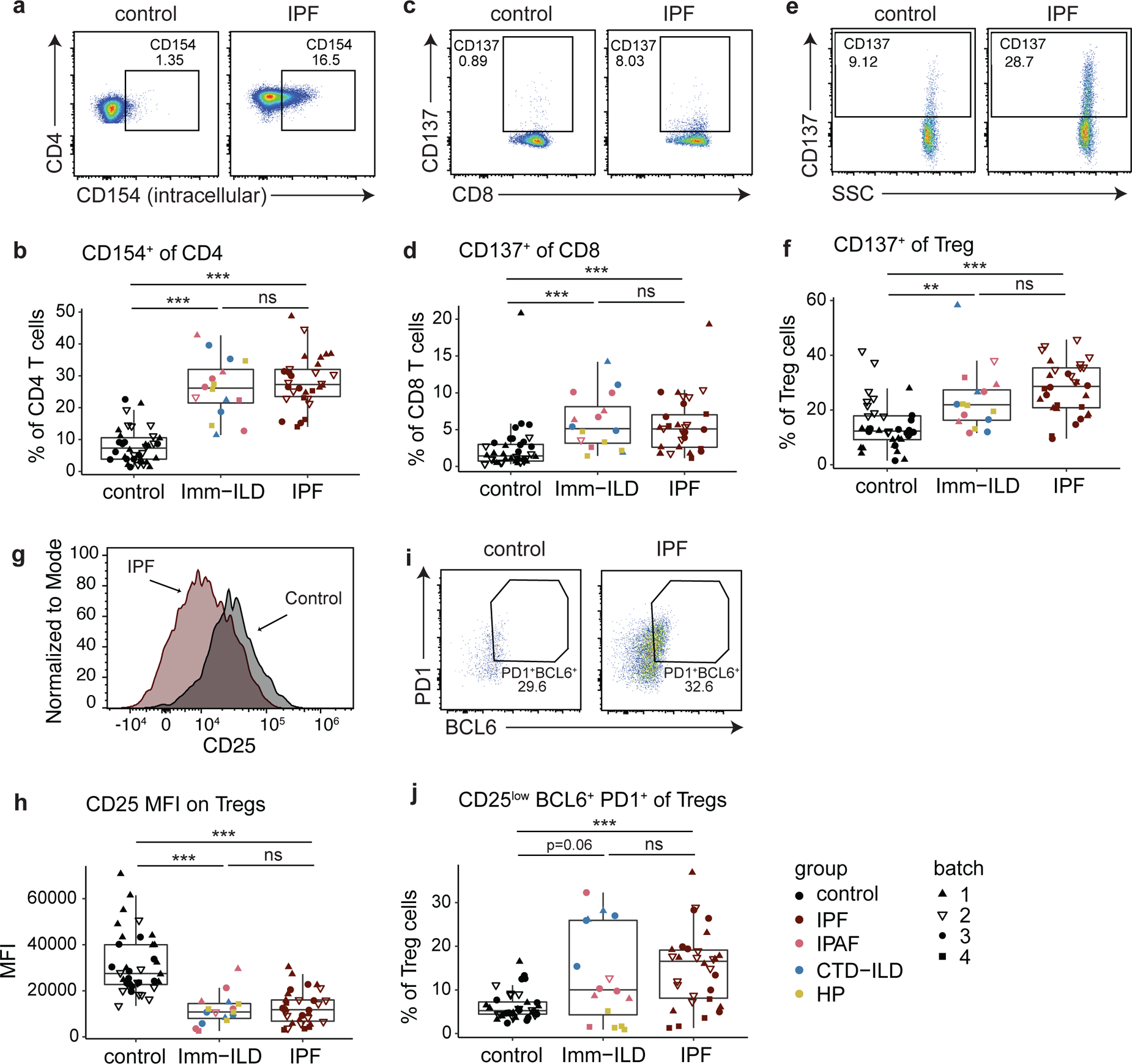
IPF and immune-mediated ILD patients have similar levels of enrichments for antigen-stimulated T cells and Treg cells in their LLNs. **a-b.** Gating strategies and quantification of frequencies of CD4 T cells expressing CD154(CD40L) in control, immune-associated ILD (Imm-ILD), and IPF LLNs. Imm-ILD group includes IPAF, CTD-ILD, and HP. **c-d.** Gating strategies and quantifications of frequencies of CD8 T cells expressing CD137(41BB). **e-f.** Gating strategies and quantifications of frequencies of Treg cells expressing CD137. **g-h.** Histograms and quantifications of median fluorescence intensity for CD25 on Treg cells. **i-j.** Gating strategies and quantifications of frequencies of Treg cells that have a follicular regulatory phenotype (CD25^low^, BCL6^+^, PD1^+^). Statistical differences in each cell subset were tested between each ILD subtypes and control LLN, by one-way ANOVA followed by Wilcoxon rank sum tests. P<0.01 (**), P<0.001 (***).

We found that the majority of Tregs in ILD patients, both IPF and immune-ILD, have low expression of CD25 (Fig. 4, G and H). Low CD25 expression is a phenotype of T follicular regulatory cells (Tfr), which can inhibit low affinity interactions between Tfh and GC B cells (Linterman et al., 2011) or prevent autoantibody production (Fu et al., 2018). Consistently, Tfr cells were significantly increased in both IPF and immune-ILD LLNs (Fig. 4, I and J), but Tfr cells accounted for only a small fraction of total Tregs. Altogether, these data suggest that enrichments of activated T cells and CD25^low^ Treg cells may be common characteristics of all fibrotic ILDs regardless of etiology.

### Germinal center responses are activated and associated with respiratory decline in ILD patients

CD154, which is highly upregulated on ILD CD4 T cells, is essential for helping B cells make antibodies. We investigated whether T and B cells involved in GC responses are also expanded in LLN of all ILD patients. Frequencies of Tfh cells and GC B cells were significantly increased in both IPF and immune-ILD LLNs compared to control LLNs, reflecting ongoing GC responses in all ILD patients (Fig. 5, A and D). Surprisingly, IPF patients had the highest amount of GC B cells, even more so than immune-ILD patients. We further interrogated GC B cells for the expression of CD10, as it has been used to define human GC B cells in lymph nodes (Clavarino et al., 2016). We found that CD10-expressing GC B cells are a significant subset of total B cells in IPF LLNs, compared to both immune-ILD patient and control LLNs (Fig. 5, E and F).

**Figure 5.**
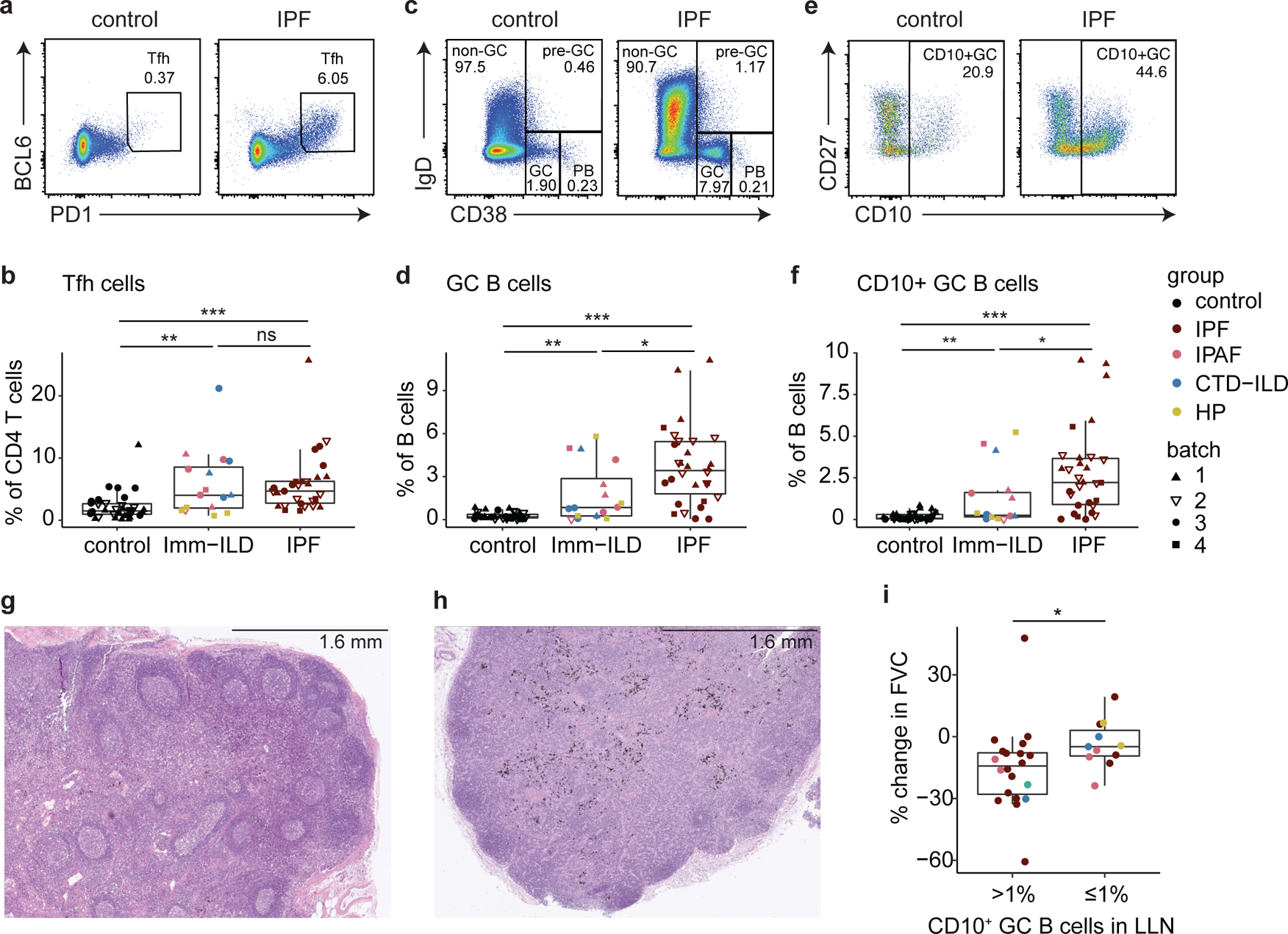
Germinal center response is activated and associated with respiratory decline in ILD patients. **a-b**. Gating strategies and frequencies of Tfh cells in control, immune-associated ILD (Imm-ILD), and IPF LLNs. **c-d.** Gating strategies and frequencies of GC B cells. **e-f.** Gating strategies and frequencies of CD10^+^ GC B cells. **g.** A representative H&E staining of LLN from end-stage IPF patients demonstrating the formation of germinal center structures **h.** A representative H&E staining of LLN from organ donor controls **i.** Comparisons of the overall percent change in forced vital capacity (FVC) over time between ILD patients who had CD10^+^ GC B cells (>1%) and ILD patients who did not have those cells (≤1%) at 1% cutoff in LLNs. P-values were obtained by Wilcoxon rank sum tests, and significance from one-way ANOVA was confirmed prior to multiple comparisons. P<0.05 (*), P<0.01 (**), P<0.001 (***).

Moreover, histology of IPF LLNs demonstrated the presence of many GC structures, while in donor control LLN, few GCs were identified (Fig. 5, G and H). Therefore, aberrant humoral immunity could be an important part of ILD pathology, especially for IPF.

Interestingly, variance in CD10^+^ GC B cell enrichment was high among ILD patients. Indeed, some patients had almost no CD10^+^ GC B cells while others had as much as 10% of CD10^+^ GC B cells in LLNs (Fig. 5 F). We hypothesized that GC B cells could have an impact on the lung and investigated if there was a relationship between the presence of an active GC response and pulmonary function. Patients with CD10^+^ GC B cell accumulation greater than 1% in their LLNs experienced worse decline in lung function as measured by the overall change in forced vital capacity (FVC) over time (Fig. 5 I). The association between CD10^+^ GC B cells and lung function was further confirmed using the mixed effects regression model (Fig. S6). The GC B cell status and time interaction coefficient suggests that patients with GC B cells experienced a further decline of FVC at about 2% per year compared to patients who did not have GC B cells (ß −1.999, 95% CI −3.304 – (−0.693), *p* < 0.003). These data suggest that activated GC responses may have negative impact on lung function in ILD patients.

### Proteome-wide screening of circulating antibodies revealed novel ILD autoantigens

Given the significant activation of adaptive immune cells and GC formation in the majority of ILD patients, we tested the hypothesis that lymphocyte activation is driven in part by endogenous antigens. To elucidate potential, we performed phage immunoprecipitation sequencing (PhIP-seq) using a library spanning the entire human proteome (Larman et al., 2011; O’Donovan et al., 2020; Vazquez et al., 2020) (Fig. 6 A). We screened serum and plasma samples collected in two medical centers, University of California San Francisco (UCSF) and University of Chicago, from 398 ILD patients at various stages of the disease, as well as 138 non-diseased control plasma samples from blood bank collections (Table 2). The most common type of ILD among this study cohort was IPF (27% of patients), followed by CTD-ILD (17%) and HP (17%). Other ILD types represented included UCTD, sarcoidosis, familial ILD, and unclassifiable ILD. From screening, we used a stringent threshold (z-score > 50) to identify antigens that are specific to ILD and not reactive in any control individual. We identified 17 candidate antigens that have not been reported elsewhere in any ILD subtype. Approximately 25% (101/398) of all ILD patients had reactivity against at least one of the 17 shared ILD peptide antigens (Fig. 6, B and C). Interestingly, each patient was positive for no more than three antigens, and the antigen specificity was heterogeneous with no apparent association to ILD subtypes (Fig. 6 D). Within the University of Chicago cohort, 51 patients were sampled more than once. We compared the first and last sample banked for each patient and found that there was no difference over time (Fig. S7).

**Figure 6.**
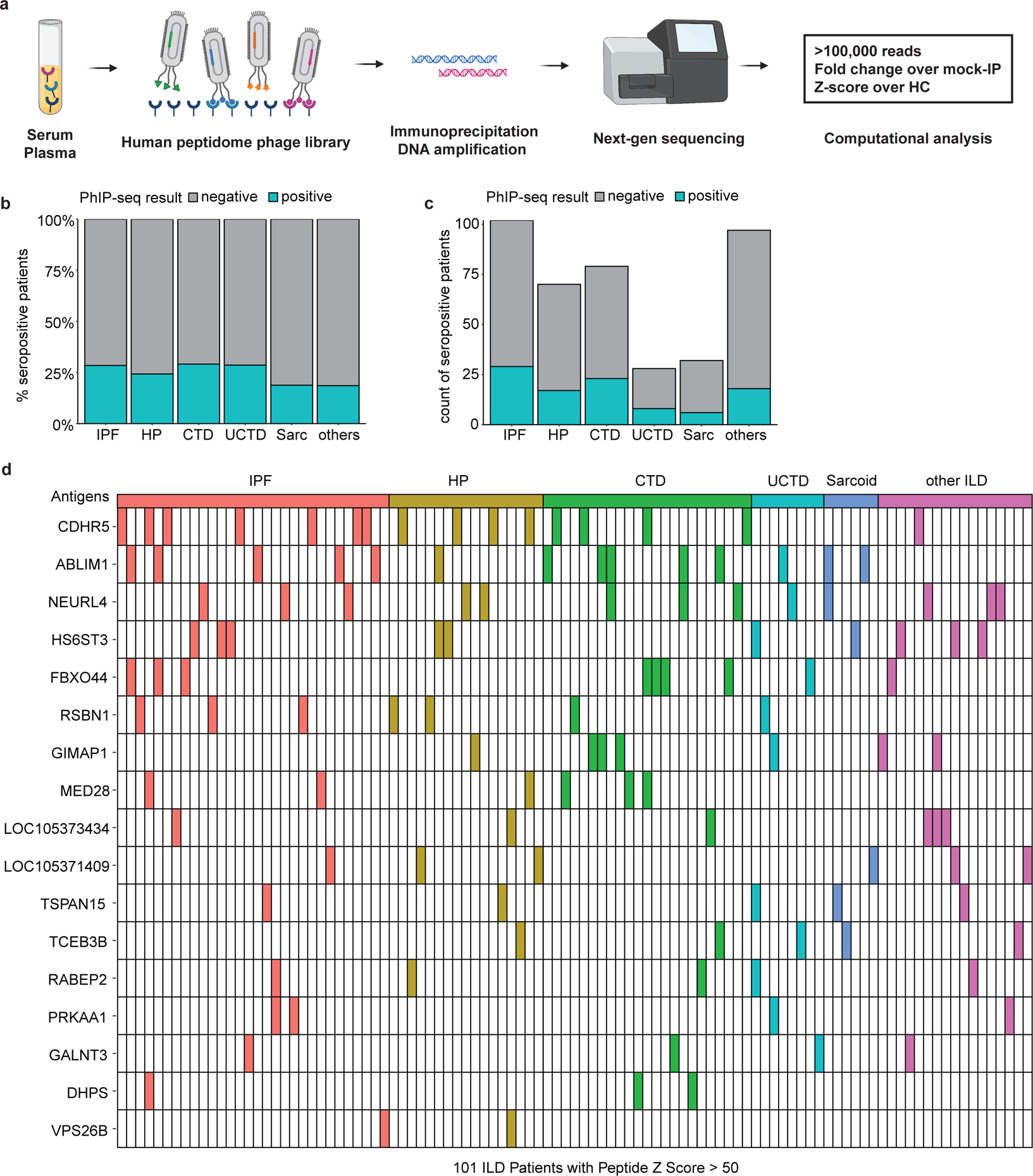
Phage-immunoprecipitation sequencing identifies novel and shared autoantigens for ILD. **a.** Using the phage library containing 700,000 unique phages, each displaying 49AA segments across the entire human proteome, a total of 398 ILD plasma/serum samples from University of Chicago and UCSF medical centers and 138 healthy control samples from the NY Blood Bank were screened. Shared autoantigens were determined where z-scores calculated relative to healthy control is above 50. **b-c.** Proportions of ILD patients who were seropositive for at least one of the shared antigens, plotted as percentage (b) and count (c). ILD individuals were grouped based on subtypes: IPF, HP, CTD-ILD (CTD), unknown CTD-ILD (UCTD; includes IPAF), sarcoidosis (Sarc), and other ILDs (others). **d.** Distribution of ILD patients who were seropositive for shared ILD antigens, ranked in the order of prevalence from top to bottom. Each column is one of the 101 ILD individuals that tested positive. None of the healthy controls had seropositivity against these antigens.

**Table 2.** Demographics of serum/plasma PhIP-seq study cohorts

| **Entire Cohort** |  |
| --- | --- |
| **Group, n** |  |
| Control (Blood Bank Samples) | 138 |
| RA (without symptoms of lung disease) | 15 |
| ILD | 398 |
| **Demographics for ILD group** |  |
| **Diagnosis, n** |  |
| Idiopathic Pulmonary Fibrosis | 102 |
| Hypersensitivity Pneumonitis | 70 |
| Connective Tissue Disease-ILD | 80 |
| Unknown Connective Tissue Disease ILD | 27 |
| Sarcoidosis | 32 |
| Others<sup>1</sup> | 87 |
| **Age, median (range)** |  |
| Idiopathic Pulmonary Fibrosis | 72 (43 – 90) |
| Hypersensitivity Pneumonitis | 67 (31 – 84) |
| Connective Tissue Disease-ILD | 61 (27 – 81) |
| Unknown Connective Tissue Disease ILD | 64 (37 – 80) |
| Sarcoidosis | 58 (29 – 75) |
| Others<sup>1</sup> | 66 (29 – 85) |
| **Sex, n** |  |
| Female | 195 |
| Male | 194 |
| Not Available | 9 |
| **Self-identified Race/Ethnicity, n** |  |
| White | 275 |
| Black | 39 |
| Hispanic, Latin(o/a/x)<sup>2</sup> | 34 |
| Asian | 28 |
| American Indian | 3 |
| Other Identity | 2 |
| Not Available | 17 |

ILD cohort consisted of a combination of n=189 from University of Chicago and n=209 from UCSF. <sup>1</sup>Others reflected a constellation of diagnoses not otherwise represented on this table and were a heterogeneous group of disorders (e.g. asbestosis, unclassifiable ILD, etc.)
<sup>2</sup>Hispanic, Latin(o/a/x) indicated individuals who did not self-identify with race but identified with ethnicity as Hispanic or Latino at UCSF and those who identified as Hispanic at University of Chicago. Individuals without known race, ethnicity, or sex information were categorized as Not Available. Data on race or ethnicity was not available for 138 subjects used as the screening reference (control) group.

### ABLIM1, a widely expressed cytoskeletal protein, is a target for autoantibody responses in many ILD patients

Using publicly available single cell RNA sequencing data and protein data, we confirmed that many of the antigenic genes or proteins are detected in lungs (not shown). Specifically, ABLIM1 is highly expressed in aberrant basaloid cells, which are transitional KRT-17^+^ epithelial cells expanded in pulmonary fibrosis patients (Neumark et al., 2020). KRT-17 is a canonical basal cell marker, which becomes upregulated in fibrotic lungs due to the expression in aberrant basaloid cells (Jaeger et al., 2022). Immunofluorescence staining of explanted IPF lungs revealed a large area of lungs positive for α-smooth muscle actin (α-SMA)-expressing fibroblasts (Shinde et al., 2017), and the presence of small airway architectures with strong cytokeratin-17 (KRT-17) expression (Fig. 7 A). Importantly, staining of IPF lungs showed strong ABLIM1 expression, which was present throughout the lungs with clear co-localization with KRT-17^+^ cells (Fig. 7 B). Immunofluorescence staining of control lungs showed a different expression pattern where expression of α-SMA and KRT-17 were restricted to large airways, but ABLIM1 was still widely expressed (Fig. S8).

**Figure 7.**
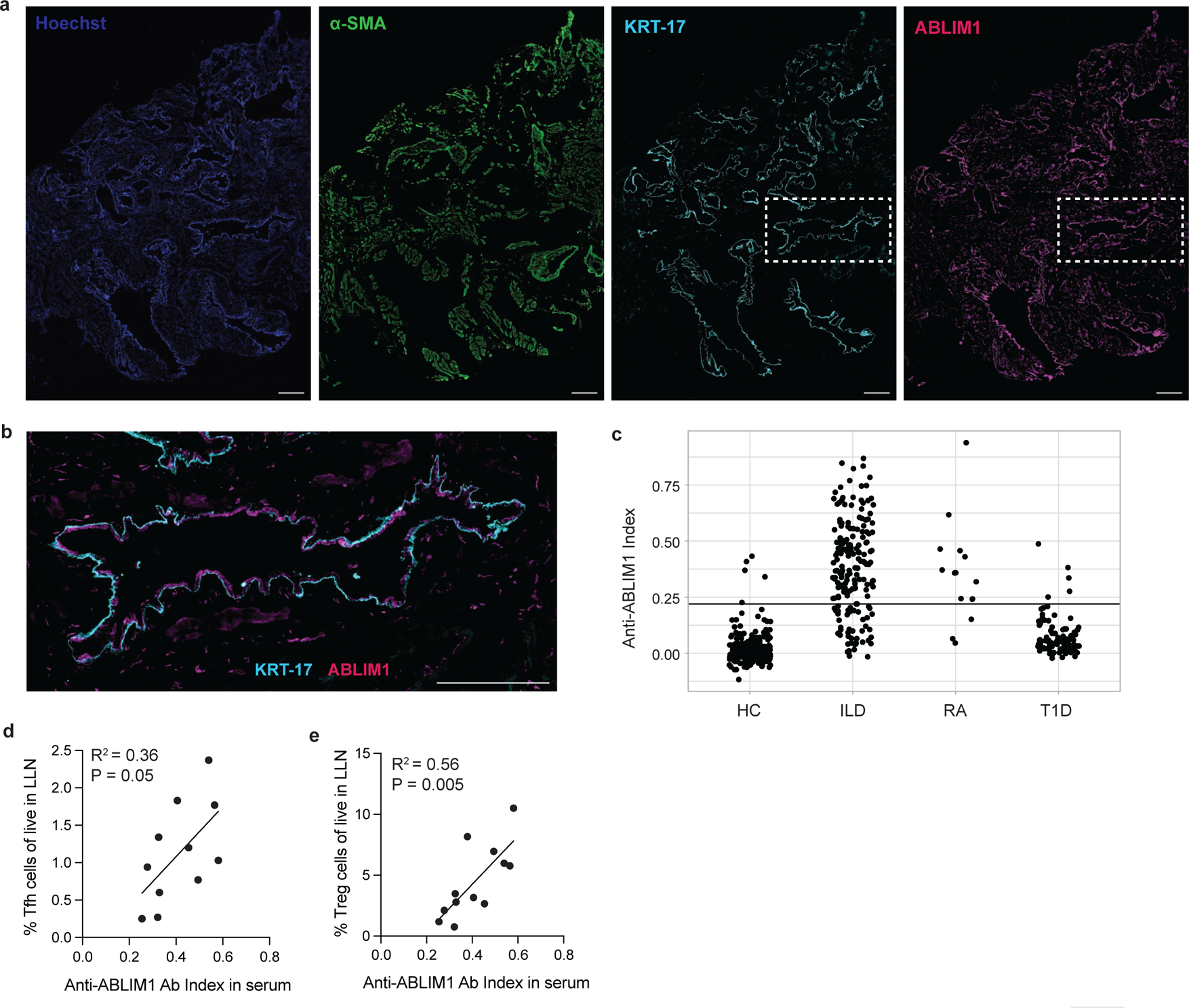
ABLIM1 is a common ILD autoantigen with high expression in fibrotic lungs. **a.** Representative immunofluorescence staining of explanted IPF lungs for Hoechst, alpha-smooth muscle actin (α-SMA), cytokeratin 17 (KRT-17), and ABLIM1. Images were taken with an 40x objective, and the scale bar is 100um. **b.** Close-up view of a region of IPF lung where there is the co-localization of ABLIM1+ cells and KRT-17+ de-differentiated airway basal cells. The scale bar is 100um. **c.** Quantification of plasma/serum antibody binding to the full-length, radiolabeled ABLIM1 protein. The horizontal line is placed at three standard deviations above the mean value for healthy controls (HC), which was used as a cutoff to determine positivity. Samples from rheumatoid arthritis (RA) patients with no symptoms of lung disease and type 1 diabetes patients (T1D) were included as comparison groups, and healthy controls (HC) are samples donated through NY Blood Bank. **d-e.** Linear regression between anti-ABLIM1 index in serum and Tfh cell (d) and Treg cell (e) proportions in ILD LLNs.

Next, we validated the binding of circulating patient antibodies to the full-length ABLIM1 protein by radioligand binding assay (RLBA) in the University of Chicago cohort. The majority of patients (142/189) tested positive for having anti-ABLIM1 antibodies in blood plasma (Fig. 7 C). To elucidate a potential immunodominant epitope on ABLIM1, we searched for hotspots based on multiple sequence alignment of the individually immunoprecipitated phage sequences. Since all known splicing isoforms were represented in our phage-displayed library, we used NCBI BLAST and determined that 21 out of 22 immunoprecipitated phage sequences aligned to either the ABLIM1 isoform a (NP_002304) or isoform j (NP001309817) (Fig. S9). Intriguingly, over 50% of the immunoprecipitated sequences aligned to a specific region of the isoform j (amino acid position 193 – 217), a proline-rich portion of the linker domain, suggesting that this could be an immunodominant epitope in ILD patents. To test whether the anti-ABLIM1 response was specific for ILD, we included a small cohort of rheumatoid arthritis (RA) patients who had no symptoms of lung disease (n = 15), and type 1 diabetes patients (n = 96) (Fig. 7 C). Interestingly, 80% of RA patients had anti-ABLIM1 autoreactivity, while only 5% of type 1 diabetes patients tested positive. Thus, we found reactivity in RA, a systemic connective tissue disease which often affects the lungs, but not in an organ-specific autoimmune disease.

Finally, we investigated whether the amount of anti-ABLIM1 antibodies in blood correlated with GC response in LLNs. We had both plasma PhIP-seq and LLN flow cytometry data from 12 individuals. We measured whether there was a linear relationship between anti-ABLIM1 index and frequency of each of the cells involved in GC responses in LLN. Although we did not find a correlation between GC B cell proportion in LLN and anti-ABLIM1 in plasma (*p* = 0.72 and R2 = 0.013), we discovered a positive linear relationship between Tfh cell proportion in LLN and anti-ABLIM1 index that was nearly significant (Fig. 7D). Moreover, we tested whether accumulation of anti-ABLIM1 antibody is related to Treg cell proportions since these regulatory cells are responsible for suppressing autoantibody production. Surprisingly, we found a strong correlation between Treg cell proportion in LLN and anti-ABLIM1 index (Fig. 7 E) suggesting that there may be unproductive suppression of GC response against ABLIM1 by Treg cells. Overall, antibody responses against common antigens expressed in fibrotic lungs suggest that these autoantigens may indeed drive adaptive immune activation that could lead to uncontrolled inflammation in lungs and lung lymph nodes.

## Discussion

Our study provides comprehensive insight into the adaptive immune landscape in the LLNs of end-stage fibrotic ILD patients who underwent lung transplantation. ILD patients’ LLNs had increased GC responses regardless of their ILD subtype. Importantly, IPF patients had immune responses as robust as patients with immune-associated fibrotic ILD, and they even had significantly more GC B cells, which associated with reduced lung function. In this active LLN environment, we hypothesized that regulatory responses would be present at low levels; however, all ILD patients had increased percentages of total and activated Tregs. As LLN T cells in the ILD patients expressed markers of recent antigen activation, we investigated whether auto-antigen responses were present in these patients. We identified serum reactivity to a novel auto-antigen, ABLIM1, and determined that anti-ABLIM1 responses positively correlated with frequencies of Tfh and Treg cells in LLNs of ILD patients. Together, these data underscore the significance of immune responses in ILD with potential roles in disease pathogenesis.

Regardless of clinical subtypes, LLN cells from all ILD patients were characterized by antigen-stimulated phenotypes exhibiting upregulation of co-stimulatory molecules. In previous work, we demonstrated that ICOS plays a protective role during lung injury in mice (Hrusch et al., 2018), and high expression of ICOS on peripheral CD4 T cells predicts favorable survival outcomes for IPF patients (Bonham et al., 2019). In the present study, we further demonstrate the importance of high ICOS expression on T cells in LLNs. ILD patients with history of hospitalization due to respiratory causes had significantly lower frequencies of ICOS-expressing CD4 T cells at the end-stage of disease in LLNs, compared to patients without a history of hospitalization. Thus, ICOS on T cells is not only predictive of survival but could also be a phenotype relevant for predicting the clinical course of the disease, such as acute exacerbation events.

In the presence of active immune responses in the LLN of ILD patients, we hypothesized there would be lower, if not the same, frequency of Tregs compared to controls. Surprisingly, we found that Tregs were elevated in all ILD patients. A significant aspect of this finding was the gating strategy, which distinguished Tregs as Foxp3^+^ CD127^−^. While previous studies on Tregs in IPF used CD25-high expression to designate Tregs, this strategy may have overlooked CD25-low expressing Tregs (Adegunsoye et al., 2016; Galati et al., 2014; Hou et al., 2017; Kotsianidis et al., 2009). We identified that a subset of CD25-low Tregs in ILD patients are Tfr cells, which regulate B cell responses by maintaining self-tolerance and downregulating ineffective GC B cell interactions (Fu et al., 2018; Linterman et al., 2011; Lu and Craft, 2021). However, Tfr responses do not seem sufficient to control auto-reactive GC B cells since we observed prevalent autoantibody responses in ILD patients. Interestingly, similar accumulation and clinical associations of CD25-low FOXP3^+^ T cells have been reported in the context of autoimmune diseases such as systemic lupus erythematosus (SLE) (Bonelli et al., 2009; El-Maraghy et al., 2018; Zhang et al., 2008), multiple sclerosis (Fransson et al., 2010), and rheumatoid arthritis(de Paz et al., 2012). Important questions beyond the scope of this study are what the origin(s) and function(s) of CD25-low FOXP3^+^ T cells in ILD are. It is possible that Tregs in ILD patients are playing a pro-fibrotic role via ICOS, CD137, or PD-1 dependent mechanisms. For example, PD-L1 ligation transforms fibroblasts into myofibroblasts, which are known to cause fibrosis (Guo et al., 2022). Thus, PD-1 expressing Tregs could be pro-fibrotic by promoting PD-L1 signaling. Future studies could investigate molecular pathways through which Treg subsets in ILD patients function and whether they contribute to fibrosis.

Enrichment of GC B cells in LLNs was associated with respiratory decline in ILD patients, suggesting a potentially pathogenic role for humoral immunity in pulmonary fibrosis progression. This is consistent with previous studies that showed a negative relationship between circulating IgA and lung function in IPF patients (Heukels et al., 2019) and a potential benefit of B cell and antibody depletions in IPF patients with acute exacerbation(Donahoe et al., 2015). Furthermore, anti-CD20 therapy using rituximab has shown to be efficacious in preserving lung function in autoimmune disease associated ILD patients (Atienza-Mateo et al., 2020). Our findings extend the B cell impact on lung function to other ILD subtypes and could support potential therapeutic benefit for treating GC B cell-high ILD patients with B cell-targeted therapies such as rituximab and anti-CD19 CAR T therapy.

The presence of germinal center responses led us to investigate whether ILD patients also had antibody responses. While there may be environmental reactivities in ILD patients, we focused on auto-antigen reactivities, as current literature suggests that many IPF patients have detectable levels of circulating autoantibodies to various proteins and cellular extracts (Feghali-Bostwick et al., 2007; Heukels et al., 2019). Despite IPF not being considered an autoimmune disease, 22-27 percent of IPF patients are seropositive for autoantibodies without meeting the full diagnostic criteria for an autoimmune or connective tissue disease (Ghang et al., 2019; Lee et al., 2013; Oldham et al., 2016). In the present study, we distinguished IPAF from IPF patients, yet still found that some autoantibodies are common across all ILD, including IPF patients without any sign of autoimmune features. We performed the first known PhIP-seq analysis on the serum and plasma of ILD patients and identified auto-reactivity to 17 candidate proteins. We highlighted ABLIM1, a protein highly-expressed in aberrant basaloid cells in fibrotic ILD lungs. Interestingly, we found that there was a break in B cell tolerance to ABLIM1 in the majority of our ILD patients, although the impact of this autoimmune response in disease pathogenesis remains unknown.

ABLIM1, which consists of an actin binding headpiece domain at carboxy-terminal and a zinc-binding LIM domain at amino-terminal, is an intracellular protein that mediates interactions between actin and cytoplasmic targets (Roof et al., 1997). While the protein is ubiquitously expressed across many tissues, some splicing isoforms have been linked to tissue-specific function or pathology of skeletal muscles (Ohsawa et al., 2015), retinal neurons (Lu et al., 2003), and osteoclasts (Narahara et al., 2018). In our PhIP-seq data, peptide epitopes for over 50% of anti-ABILM1+ ILD patients mapped to a region that only exists in the isoform j (NP_001309817), which is a shorter isoform (455AA) compared to others. Further study is needed to determine exon-splicing profiles of ABLIM1 expressed in fibrotic lungs. Interestingly, we did not find a correlation between anti-ABLIM1 index and ILD subtypes, as distributions of ILD subtypes were nearly identical between the full cohort and the seropositive subset. This is consistent with our findings from the flow cytometry analysis of LLN, which revealed lymphocyte activation including germinal center responses across all ILD types.

Antibodies to ABLIM1 were also detected in RA patients, but not in type 1 diabetes patients, suggesting that the break in tolerance against ABLIM1 might be due to some shared pathology between ILD and RA. In fact, pulmonary involvement is one of the common extra-articular manifestation of RA (Yunt and Solomon, 2015), and even patients without symptoms of lung disease can have radiographic lung abnormalities (Bilgici et al., 2005). Though investigation of a larger RA cohort is needed, it is possible that anti-ABLIM1 responses are specific to lung pathologies. The lack of response in type 1 diabetes patients further supports that ILD-specific mechanisms rather than chronic inflammation likely contribute to autoantibody response.

While PhIP-seq screening covers the entire human proteome, this method is limited to linear peptides which would exclude complex epitopes and post-translational modifications of an entire protein. In fact, anti-ABLIM1 prevalence turned out to be greater using RLBA of full-length protein compared to PhIP-seq analysis, suggesting strong immunogenicity of 3-dimensional epitopes. In addition, smoking, a risk factor in ILD, induces a post-translational modification called citrullination (Makrygiannakis et al., 2008). Citrullination can result in major structural changes including charge shift, hydrogen bond formation, and protein denaturation (Alghamdi et al., 2019). Thus, PhIP-seq may have missed reactivity of citrullinated peptides. These caveats of PhIP-seq may explain why we did not detect previously published auto-antigen reactivities.

The initial trigger leading to enlarged lymph nodes remains an outstanding question. Although we provide compelling data on auto-antigen reactivities in ILD, it is entirely possible that there are also environmental antigens or even inhaled atmospheric particulate matter (Ural et al., 2022) participating in the activation of immune responses in the LLN. Further, acute exacerbations in ILD patients accelerate lung function decline, and thus infectious pathogens, which often lead to acute exacerbations, may contribute to immune activation and antibody production. Finally, although we had a large number of explanted LLN samples, we did not have enough power to include medication history as a variable. Thus, the relative increase in GC B cell enrichment in LLNs of IPF patients compared to immune-ILD patients could reflect differences in immunomodulatory treatments between the two groups.

Together, these data provide an interesting new avenue for research on human ILD. We provide evidence that there is an antigen response occurring in ILD patients and importantly, there is a commonality amongst IPF and immune-associated ILDs at the end-stage of disease. With these data, we can hypothesize that auto-antigens may be driving germinal center responses and activation of lymphocytes that are contributing to pathogenesis of disease. However, autoantibody production could be either a driver or byproduct of the disease. Although our analysis of paired samples nullified a hypothesis that there is an increasing autoantibody response over time, ILD patients become seropositive at various stages of the disease. In contrast to the view that end-stage fibrotic ILD is a burnt-out fibrosis without immune involvement, our work implies that inflammation and autoimmunity remain ongoing even at time of lung transplantation. In addition, our findings suggest that a break in humoral tolerance is a widespread phenomenon and that there may be uncontrolled immune responses against common autoantigens in all types of fibrotic ILDs, even in IPF. Future studies on clinical phenotypes and prognosis of patients who are seropositive for ILD autoantigens could potentially define novel, immediately relevant endotypes.

## Materials and Methods

### Collection of human tissues

LLN cells and lung tissue sections were collected from ILD patients who underwent lung transplantation at the University of Chicago between August 2013 and December 2019. Control lung and lymph node samples were obtained from the Illinois Gift of Hope Organ & Tissue Donor Network. Lungs were received to the lab and processed within 48 hours of clamp time. LLN were dissected from around the trachea and bronchi of lungs from the Gift of Hope, and from the lung parenchyma of ILD explanted lungs. LLNs were cut into small pieces (about 5mm^3^) and pressed through sterile nytex to release cells from the outer capsule layer. Viable lymphocytes were purified through centrifuging the cells after being layered onto Histopaque-1077 medium (Sigma Aldrich, St Louis, MO) at 1750rpm for 20 min without brakes. The purified lymphocytes were resuspended in freezing medium consisting of 90% fetal calf serum (X&Y Cell Culture, Kansas City, MO) and 10% DMSO (Sigma-Aldrich, St Louis, MO), aliquoted into cryovials and preserved in liquid nitrogen until use.

Lung parenchyma tissue sections (about 1 - 1.5 cm^2^) were dissected from the right lower lobes of lungs received from the Gift of Hope, and from central (less fibrotic) or peripheral (more fibrotic) regions of the right and left lung lobes from IPF lung transplant patients. Lung tissues were placed into cryomolds, embedded in OCT Compound (OCT 4583, Fisher HealthCare, Waltham, MA), frozen, and stored at −80°C until sectioning. For histology of LLN sections, formalin-fixed, paraffin-embedded (FFPE) blocks were prepared. Tissues were placed in individual cassettes and fixed in 10% formalin (Azer Scientific, Morgantown, PA). Fixed samples were washed in 70% ethanol and water and were embedded in paraffin wax by the Human Tissue Resource Center at University of Chicago.

Serum and plasma samples for PhIP-seq analysis were obtained from patients at University of Chicago and UCSF medical centers. Red-capped Vacutainer blood collection tubes (BD, Franklin Lakes, NJ) were spun at 1500 rpm for 10 minutes at 4°C. The top layer of serum was removed and stored in aliquots at −80°C. Control plasma samples were obtained from blood banks in San Francisco and New York.

The study is approved by Institutional Review Boards (University of Chicago IRB 14163A & IRB 14514A, UCSF IRB 10-02467 & IRB 10-01592), and all ILD patients included in the study gave informed consent. Samples from organ donors are IRB exempt as they are deceased upon donation.

### Collection of clinical data

Diagnosis of ILD was established at University of Chicago based on multidisciplinary discussion of pulmonologists, dedicated chest radiologists, rheumatologists, and a thoracic pathologists in concordance with the American Thoracic Society/European Respiratory Society criteria (Fischer et al., 2015; Raghu et al., 2018; Raghu et al., 2020). For PhIP-seq study, two authors from UCSF and University of Chicago (V.U. and C.T.L.) collapsed diagnostic categorization of ILD to shared data categories as detailed in (Upadhyay et al., 2023). RA patients’ sera used in RLBA was patients who had no symptoms of lung disease. From consented patients, race, ethnicity, date of birth, pulmonary function test results, hospitalization records, and lymphadenopathy information were collected. Lymphadenopathy larger than 10 mm in diameter was noted in the radiology reports of high-resolution chest computerized tomography scan. Information on lymphadenopathy status and frequency and cause of hospitalization of patients that are included in this study were collected by pulmonologists through patient chart review. Patient information was de-identified and maintained with a randomly generated unique identifier specific to each subject. Demographic information and donor clinical history were provided by the Gift of Hope. Information was pooled from the donors’ medical chart and from family-reported social and medical histories, including smoking status, drug use, and any known airway disease.

### Cryorecovery of lymphocytes from lung lymph nodes

Cryopreserved LLN samples from explanted ILD lungs and non-ILD organ donor lungs were thawed in 37C water bath with constant swirling. The cells were resuspended in 4ml of HBSS containing 2% FCS (X&Y) and 10ug/ml DNase I (LS002138, Worthington Biochemical, Lakewood, NJ) and layered on top of 3ml of Histopaque-1077 (Sigma). After spinning at 1750rpm for 20min with no brake, live lymphocytes were collected from the interphase and washed once with 2% FCS in RPMI.

### Flow cytometry analysis

0.5 to 1.0 x 10^6^ cells from each sample were aliquoted into 5ml round bottom tubes or 96-well U-bottom plates for antibody staining. Cells were washed with PBS and stained for viability using Zombie NIR Fixable Viability stain (1:2500, BioLegend 423106) in PBS for 20 min at room temperature in dark. Cells were washed with FACS buffer, blocked with Human TrueStain FcX (1:200, Biolegend 422302), and stained with the surface antibody cocktail in FACS buffer for 30 min at 4C in dark. All antibodies and dilutions used for staining are listed in Table S1 and Table S2.

For intracellular staining, cells were fixed and permeabilized using the Foxp3/Transcription Factor Staining Buffer Set (eBioscience 00-5523-00). To block non-specific binding, 10ul of FBS was added to the cells in the residual volume of perm buffer and incubated for 5 min. Without washing, the intracellular antibody cocktail was added to the cells and stained for overnight at 4C. After the staining, the cells were washed twice with perm buffer, once with FACS buffer, and resuspended in FACS buffer for analysis. Antibody-stained cells were analyzed on an Aurora (Cytek) and the data analysis was performed using FlowJo (BD). Dimensionality reduction and hierarchical clustering analyses were performed using R functions and packages including ComplexHeatmap.

### Phage immunoprecipitation-sequencing

PhIP-seq was performed as previously described to screen circulating patient antibodies against approximately 700,000 different linear peptide segments, each at 49 amino acids in length, spanning the entire human proteome (Larman et al., 2011; O’Donovan et al., 2020; Vazquez et al., 2020). Based on the next-generation sequencing results, patient antibody-bound peptide sequence counts were quantified as fold change (FC) over mock immunoprecipitated (beads, no serum) sequence counts. To control for noise, we only considered sequences with greater than 10 FC over negative controls for downstream analysis. Next, to distinguish disease-relevant antigens, we normalized the FC values against the mean FC value of non-diseased controls for each given sequence as z-scores. We used a z-score threshold of values greater than 50 to identify antigens that are strongly enriched in ILD patients over non-diseased controls. Finally, peptide sequences were aligned to genes, and common antigens are defined as positivity in 2 or more individuals.

### Radioligand binding assay

Full length cDNA sequences for ABLIM1 (NM_001322888) led by the Kozak sequence (GCCACC) were cloned into pTNT vector (Promega L5610), and the constructs were validated using Sanger sequencing. Using the rabbit reticulocyte lysate system (Promega TNT Quick Coupled Transcription/Translation System), proteins were translated under the control of T7 promoter with 35S labels in vitro. Proteins were purified by passing through Nap-5 column (GE 17-0853-01) and quantified using the MicroBeta Trilux liquid scintillation counter (PerkinElmer). Proteins were confirmed based on size after running on SDS-PAGE gels.

To quantify binding of patient antibodies to radiolabeled protein antigens, 5ul of plasma or serum samples were incubated with 5,000 – 20,000 counts of labeled protein overnight. On the next day, the antibody complex was immunoprecipitated using 4:1 protein A:G agarose beads (GE) on 96-well plates with a PVDF filter membrane. After washing the plate 4 times using a vacuum apparatus and air drying for 30min, the ionizing radiation from immunoprecipitated antibody-ligand complex was quantified using the scintillation counter. Every plate included A:G beads only (negative controls) and commercial antibody (positive controls) in triplicates. Commercial anti-ABLIM1 (Novus Biologicals NBP198472) was used as the positive control. The antibody index for each plasma sample was calculated by computing (sample value – mean value negative control value)/(mean positive control value – mean negative control value). Experimental samples with values 3 standard deviations above the mean of healthy controls for each antigen were considered positive.

### Immunofluorescence

Explanted lung pieces from ILD patients and organ donors were freshly frozen in optimal cutting temperature compound (OCT 4583) and stored at −80C. Tissue blocks were cut into 8μm-thin serial slices and mounted onto slides. Tissue sections were fixed with 4% PFA for 10 min, permeabilized with 0.1% NP-40 for 10 min, quenched with freshly made 0.1mg/ml NaBH_4_ for 5min, and blocked with 10% normal donkey serum and Human TrueStain FcX (1:200, Biolegend 422302) for 30 min. Slides were stained with primary antibodies overnight at 4C using rabbit anti-human ABLIM1 (1:200, Proteintech 15129-1-AP) and mouse anti-human cytokeratin 17 (1:100, Invitrogen MA 1-06325) diluted in PBS with 5% normal donkey serum. Tissues were washed and stained with secondary antibodies Alexa Fluor 647-donkey anti-rabbit (1:1000, Invitrogen A31573) and Alexa Fluor 568-donkey anti-mouse (1:1000, Invitrogen A10037) in PBS with 5% normal donkey serum for 1 hour at room temperature. After washing, sections were blocked with 10% mouse serum for 30 min, and then stained using a directly conjugated Alexa Fluor 488-anti-alpha smooth muscle actin monoclonal antibody (1:250, Invitrogen 53-9760-82) for 2 hours at room temperature. The slides were washed and nuclear stained with Hoechst 33342 (1:1000, Invitrogen H3570) for 30 min. Finally, the slides were mounted with ProLong Gold Antifade Mountant (Invitrogen P36934) and cured for 24 hours at room temperature. For every sample, an adjacent section was stained with only secondary antibodies as a control. Slides were imaged by the University of Chicago Integrated Light Microscopy Core, using an Olympus VS200 Slideview Research Slide Scanner with a 40X objective. The images were analyzed using QuPath (Bankhead et al., 2017) and ImageJ (Schneider et al., 2012) softwares.

### Statistical analysis

Statistical comparisons of specific cellular frequencies between disease groups and controls were achieved by non-parametric Mann Whitney Wilcoxon Test and also by one-way ANOVA with multiple comparisons when there were three groups. To test associations between having an ILD and protein expression (MFI from flow cytometry data), a binomial logistic regression analysis was performed for each flow parameter using the glm() function in R, where: disease state = β0 + β1 ∗ MFI + β2 ∗ batch. Likelihood ratio test (LRT) for β1 was performed to test the significance of MFI of each flow parameter, and p-values were adjusted using the Benjamini-Hochberg (BH) method via the p.adjust() function in R. To test relationships between having an ILD and the enrichment of immune cell populations in LLN, a generalized linear model for each population was built with a Gaussian distribution, where: ln(cell frequency)/(1 − cell frequency) = β0 + β1 ∗ disease state + β2 ∗ batch. The p-value for β1 was calculated by LRT and adjusted for multiple testing by BH method. To test whether the enrichment of germinal center B cells in LLN is associated with changes in pulmonary function over time, a mixed effects model was built using STATA (StataCorp, College Station, TX). The status of germinal center B cell enrichment in each patient was converted into a binary response (GCB status: yes or no) using a cutoff of 1% of live cells in LLN. Longitudinally collected forced vital capacity (FVC) from ILD patients was used as a pulmonary function measurement. The length of time between each pulmonary function test was calculated in years and included in the model, where: E[FVC|GCB status, year] = β0 + β1 ∗ GCB status + β2 ∗ year + β3 ∗ GCB status * year. The coefficient and p-value for the interaction term between GCB status and year were reported to determine how much FVC change per year differs between GCB-positive and GCB-negative patients. Simple linear regressions were performed to test relationships between the antibody index against ABLIM1 and immune cell proportions in LLN. The Goodness of fit (R-squared value) and p-value were obtained using GraphPad Prism (GraphPad Software, San Diego, CA).

## Data Availability

All data produced in the present study are available upon reasonable request to the authors.

https://derisilab.ucsf.edu/

## Acknowledgments

We thank the Flow Cytometry Core Facility at the University of Chicago. We thank Catherine S. Bonham and Rebecca Voogt for contributing to the development of the human sample bank and the banking of human samples.

This work was supported by National Institute of Health grants: U19 AI162310 (A.I. Sperling), R01 HL13079 (I. Noth), T32 AI007090-44 (Y. Yoon), K08HL165106 (V. Upadhyay), R01AI68299 (A.K. Shum), U19 AI082724 (M.R. Clark), T32 HL007605 (K.C. Selvan), K23 HL146942 (A. Adegunsoye), F31 HL156659 (M.K. Hollinger), The American Heart Association (T.E. Velez), The Pulmonary Fibrosis Foundation (C.T. Lee), The Nina Ireland Program for Lung Health at UCSF (P.J. Wolters), The UCSF Clinical Translational Sciences Institute (V. Upadhyay).

## Author contributions

Y. Yoon, T.E. Velez, V. Upadhyay, A.K. Shum, A.I. Sperling were involved in all aspects of this study, including conceiving the project, designing and performing experiments, data analyses and interpretation, and manuscript writing. S.E. Vazquez, M. Anderson, J.L. DeRisi contributed to the Phip-seq studies and analysis. C.S. Law, K.M. Blaine, M.K Hollinger, D.C. Decker, provided experimental assistance and contributed to the manuscript. V. Upadhyay, C.T. Lee, K.C. Selvan, M.R. Clark, M.E. Strek, R.D. Guzy, A. Adegunsoye, I. Noth, P. J. Wolters, provided clinical samples, clinical data, and supervision. A.K. Shum, A.I. Sperling, J.L. DeRisi, and M. Anderson provided financial and experimental resources. A.I. Sperling provided overall supervision of the study.

