## Supplemental Figures for "Antigenic responses are hallmarks of fibrotic interstitial lung diseases independent of underlying etiologies"

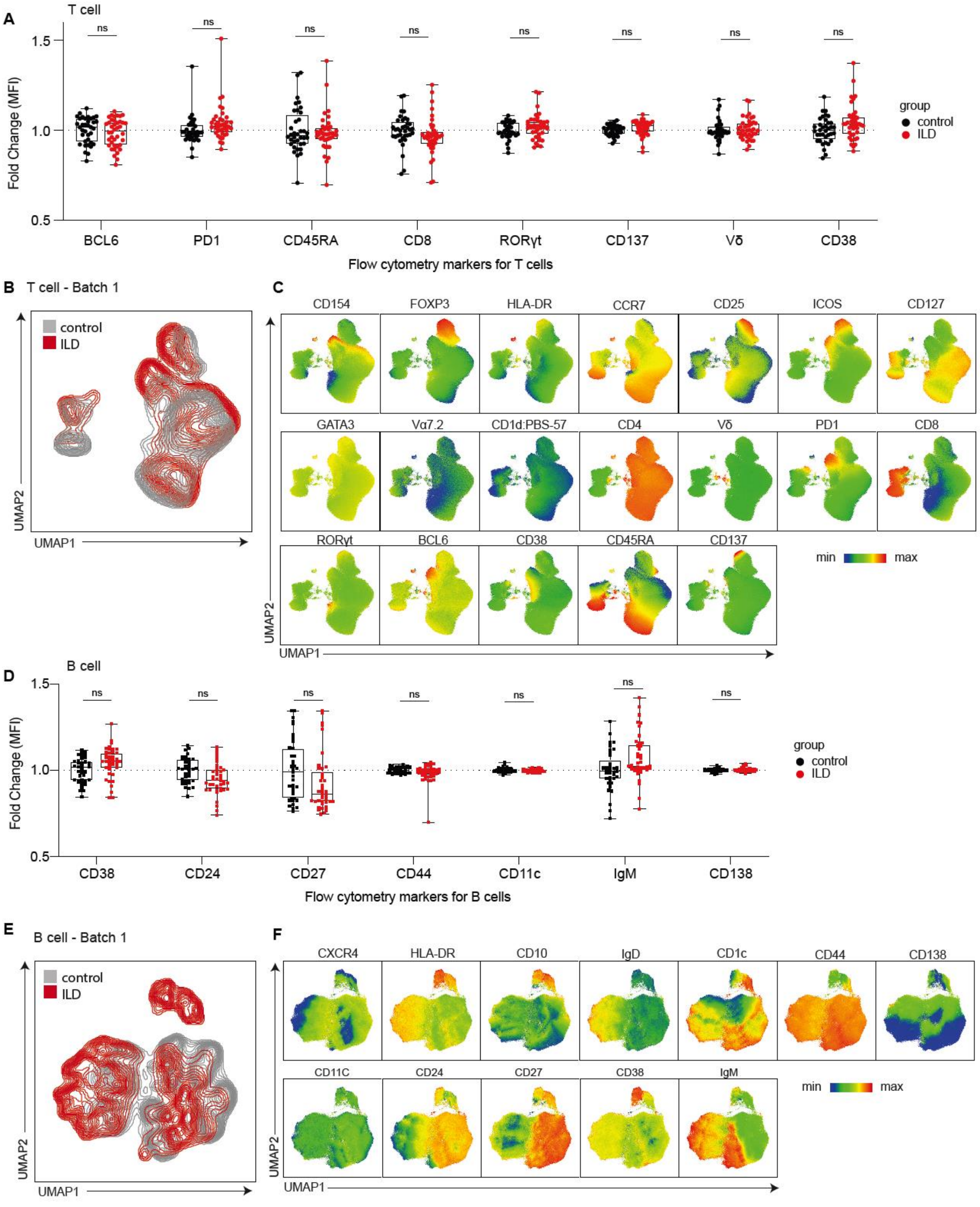


Figure S1. **T cell and B cell landscapes in LLNs from ILD patients compared with matched controls. (A)** Box plots comparing the median fluorescence intensity of individual T cell markers based on all samples in batches 1-3. Fold change was calculated relative to the mean values of control samples, separately for each batch. **(B)** Representative UMAP of T cells. Each group (control vs ILD) contains 130,000 T cells concatenated from 13 individuals (10,000 cells per individual) in batch 1. **(C)** Fluorescence intensity of each T cell parameter overlaid on the UMAP plot. **(D)** Box plots comparing the median fluorescence intensity of individual B cell markers based on all samples in batches 1-3. Fold change was calculated relative to the mean values of control samples, separately for each batch. **(E)** Representative UMAP of B cells. Each group (control vs ILD) contains 130,000 B cells concatenated from 13 individuals (10,000 cells per individual) in batch 1. **(F)** Fluorescence intensity of each B cell parameter overlaid on the UMAP plot.

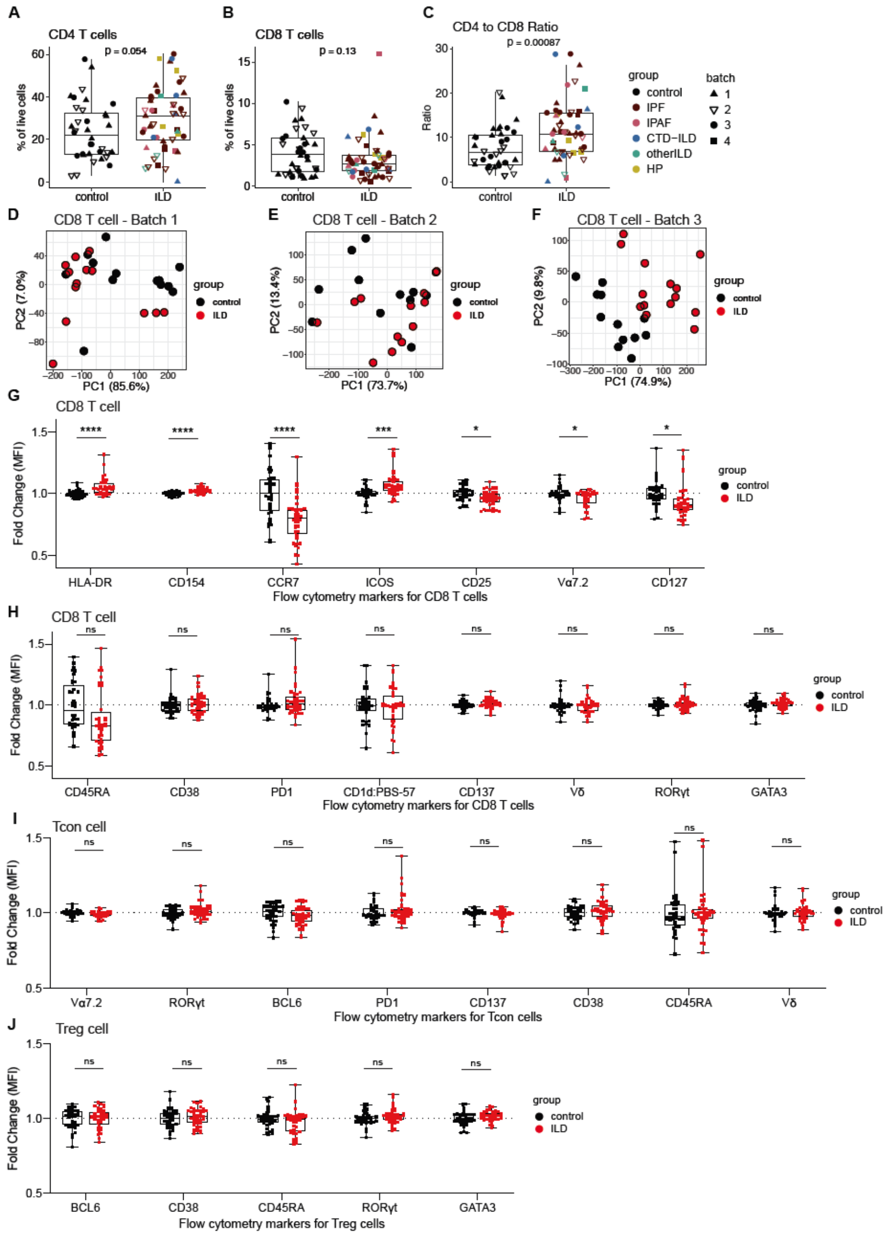


Figure S2. **T cell subpopulations in LLNs from ILD patients compared with matched controls. (A)** Comparison of CD4 T cell frequencies in control and ILD LLNs. **(B)** Comparison of CD8 T cell frequencies in control and ILD LLNs. **c.** Comparison of CD4 to CD8 T cell ratio in control and ILD LLNs. **(D-F)** PCA of CD8 T cells based on fluorescence intensities of 15 parameters: CD154, HLA-DR, CCR7, CD25, ICOS, CD127, GATA3, Vα7.2, PBS-57 loaded CD1d tetramer, Vδ, PD1, RORγt, CD38, CD45RA and CD137. An analysis was run with 1,600 cells, 669 cells, and 1,100 cells for batch1, batch2, and batch 3, respectively. **(G-H)** Box plots comparing the median fluorescence intensity of individual CD8 T cell markers based on all samples in batches 1-3. **i.** Box plots comparing the median fluorescence intensity of individual Tcon markers based on all samples in batches 1-3. **j.** Box plots comparing the median fluorescence intensity of individual Treg markers based on all samples in batches 1-3. For all MFI comparisons, fold change was calculated relative to the mean values of control samples, separately for each batch. Parameters that are significantly associated with ILD based on a likelihood-ratio test on each logistic regression coefficient are shown. P-values were adjusted by Benjamini-Hochberg correction. $P<0.05$ (*), $P<0.01$ (**), $P<0.001$ (***), $P<0.0001$ (****).

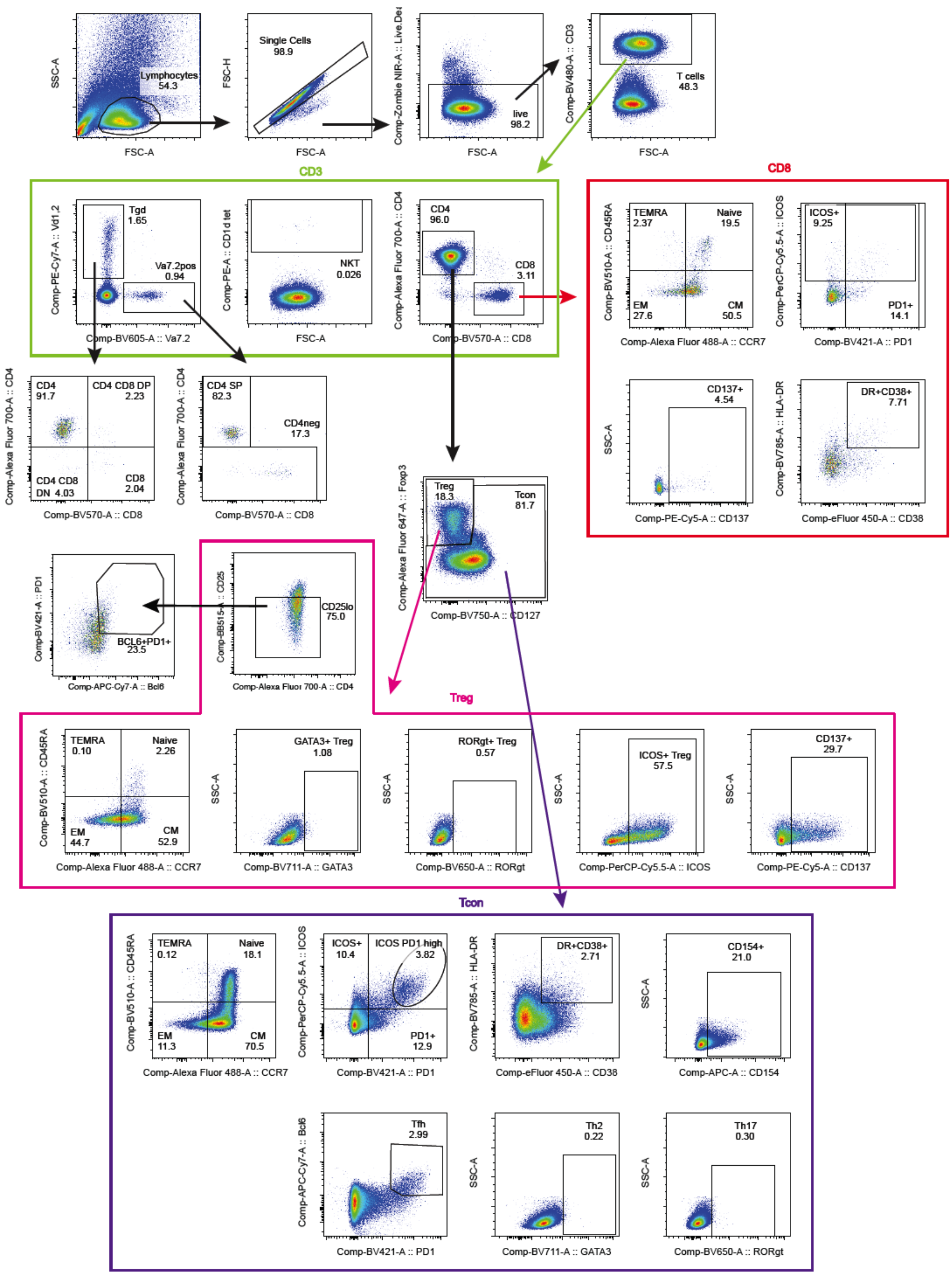


Figure S3. **Gating strategies for T cell populations.**

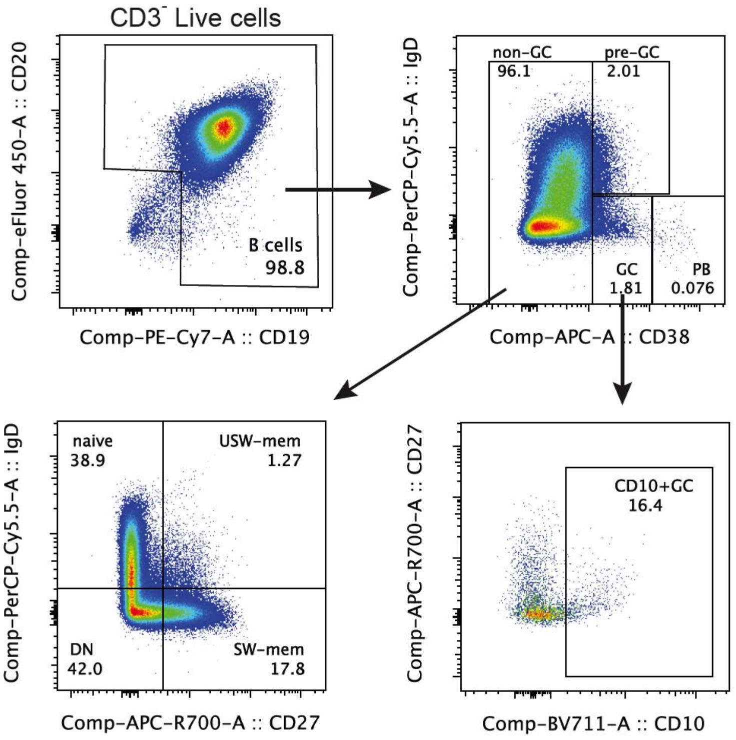


Figure S4. **Gating strategies for B cell populations.**

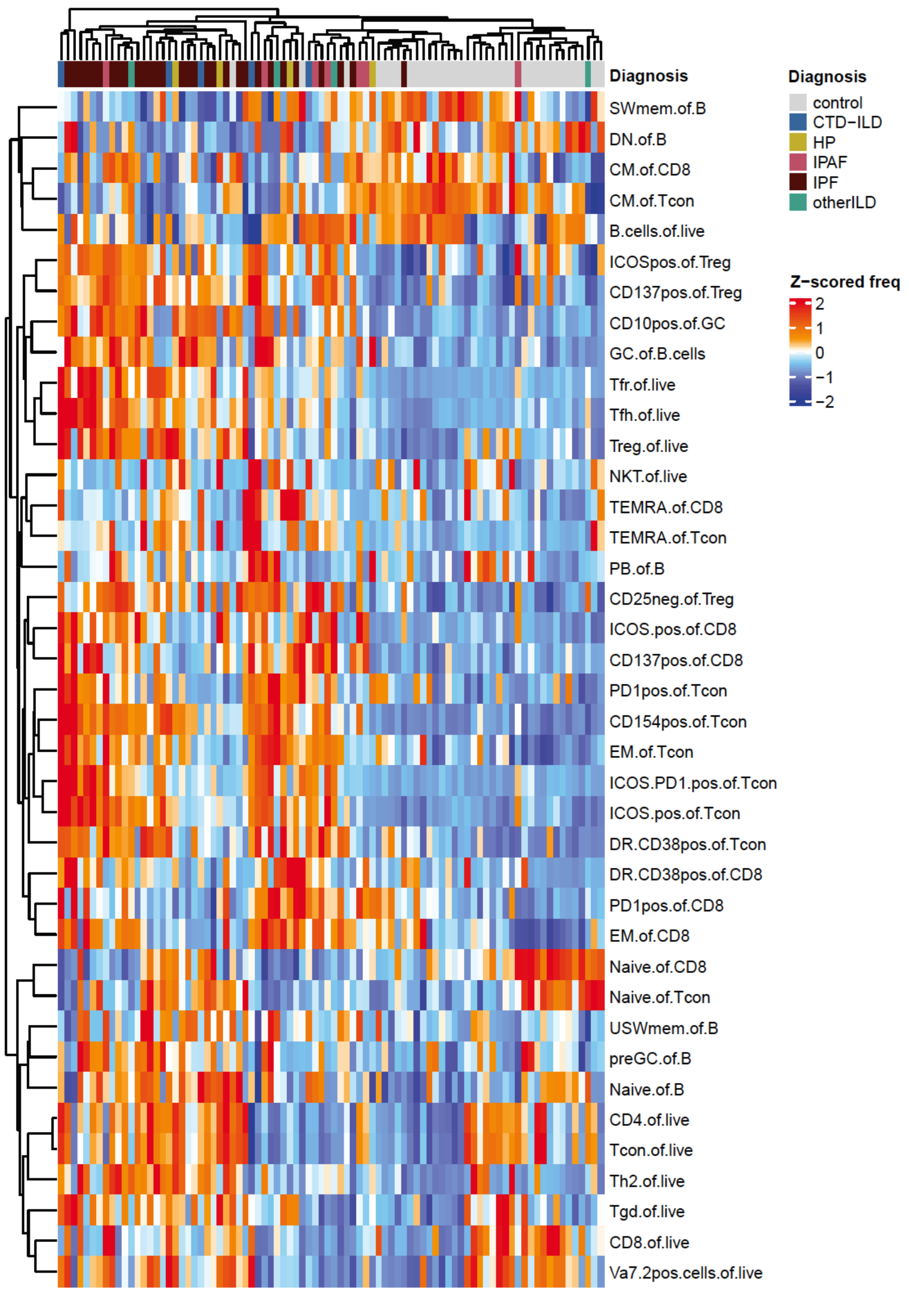


Figure S5. **Heatmap with unsupervised clustering based on immune cell abundance.** Heatmap visualization of a hierarchical clustering of control and ILD individuals based on z-scored frequencies of 39 immune cell populations.

```
Mixed-effects ML regression                     Number of obs     =        274
Group variable: sampleID                        Number of groups  =         31

                                                Obs per group:
                                                              min =          2
                                                              avg =        8.8
                                                              max =         37

                                                Wald chi2(3)      =      54.83
Log likelihood =  -972.3637                     Prob > chi2       =     0.0000

------------------------------------------------------------------------------
         fvc |      Coef.   Std. Err.      z    P>|z|     [95% Conf. Interval]
-------------+----------------------------------------------------------------
        year |  -1.453615   .3754472    -3.87   0.000    -2.189479   -.7177524
  GCB_status |   6.396411   5.580307     1.15   0.252     -4.54079    17.33361
    GCB*year |   -1.99864   .6660458    -3.00   0.003    -3.304066   -.6932146
       _cons |   55.72422   3.998668    13.94   0.000     47.88697    63.56146
------------------------------------------------------------------------------

------------------------------------------------------------------------------
  Random-effects Parameters  |   Estimate   Std. Err.     [95% Conf. Interval]
-----------------------------+------------------------------------------------
mrn: Identity                |
                  var(_cons) |   224.9574   59.48363      133.9751    377.7257
-----------------------------+------------------------------------------------
               var(Residual) |   47.42213   4.303841      39.69441    56.65427
------------------------------------------------------------------------------
LR test vs. linear model: chibar2(01) = 340.84        Prob >= chibar2 = 0.0000
```

Figure S6. **Relationship between the lung function changes over time and GC B cell status in explanted LLNs.** Mixed-effects regression modeling with a random intercept was used to analyze changes in forced vital capacity (FVC) over time (year) by germinal center B cell status (GCB_status) in explanted LLNs.

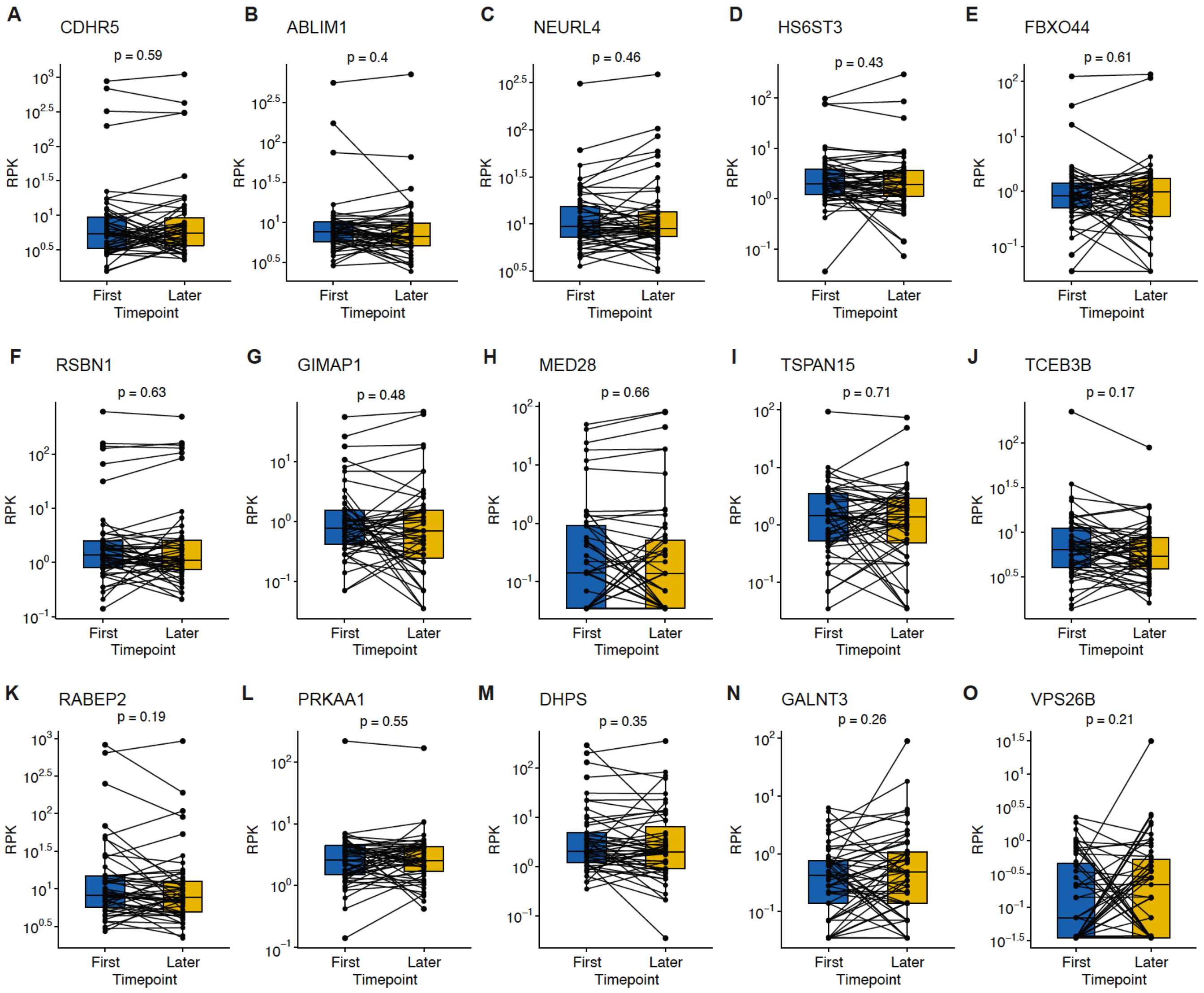


Figure S7. **Longitudinal analysis of autoantibody amounts in ILD plasma. (A-O)** Autoantibody signals based on PhIP-seq results (RPK: reads per 100k) were compared between longitudinally paired plasma samples from ILD patients (n=51). Intervals between the first and later time-points range between 98 to 2952 days. Paired Wilcoxon p-values are displayed for each of the 15 ILD antigens.

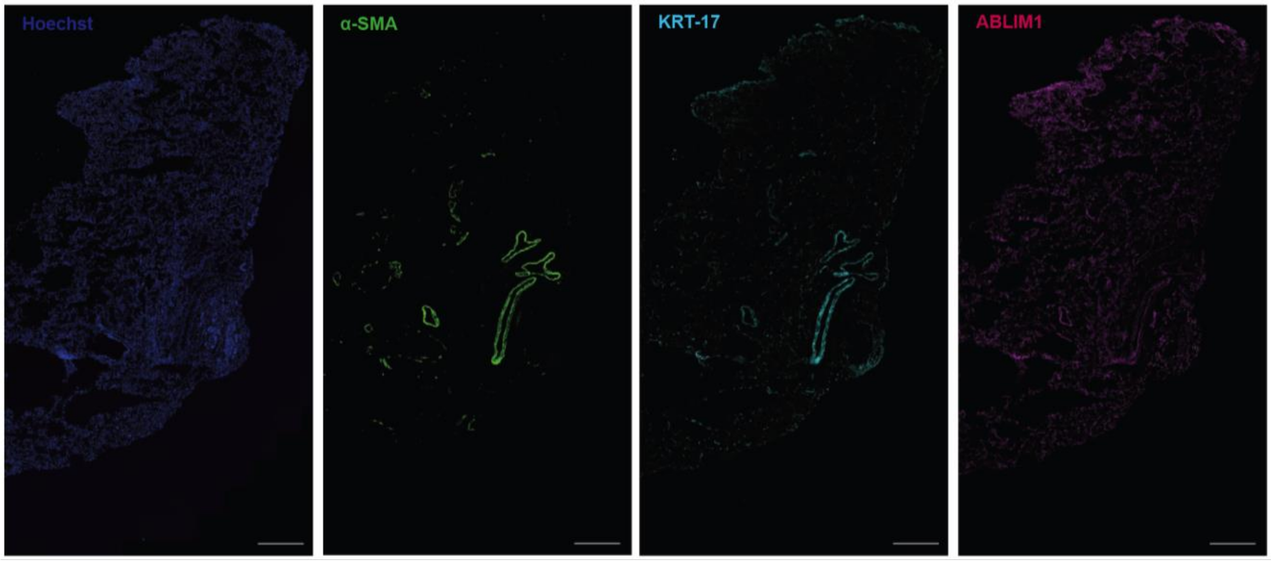


Figure S8. **Immunofluorescence analysis of ABLIM1 in control lungs.** Representative immunofluorescence staining of organ donor control lungs for Hoechst, alpha-smooth muscle actin (α-SMA), cytokeratin 17 (KRT-17), and ABLIM1. Images were taken with a 40x objective, and the scale bar is 100um.

```
Isoform a      | 1 MPAFLGLKCLGKLCSSEKSKVTSSERTSARGSNRKRLIVEDRRVSGTSFTAHRRATITHLLYLCPKDYCPRGRV 74
Isoform j      |   --------------------------------------------------------------------------
Epitope (n=3)  |   MPAFLGLKCLGKLCSSEKSKVTSSERTSARGSNRKRLIVEDRRVSGTSF-------------------------
Epitope (n=1)  |   ------------------------ERTSARGSNRKRLIVEDRRVSGTSFTAHRRATITHLLYLCPKDYCPRGR-

Isoform a      | 440 QDVRDRMIHRSTSQGSINSPVYSRHSYTPTTSRSPQHFH--------------
Isoform j      | 124 QDVRDRMIHRSTSQGSINSPVYSRHSYTPTTSRSPQHFH--------------
Epitope (n=1)  |     -DVRDRMIHRSTSQGSINSPVYSRHSYTPTTSRSPQHFHRPDQGINIYRK---

Isoform a      |     -------------------------------------GNEPSSGRNSPLPYRPDSRPLTPTYAQAPKHFHVPDQGINIYRKPPIYKQH 531
Isoform j      | 168 SPGVQRLSYLRTSSLSPTHSDSRPNPPFRHHFIPHIKGNEPSSGRNSPLPYRPDSRPLTPTYAQAPKHFHVPDQGINIYRKPPIYKQH 255
Epitope (n=5)  |     -PGVQRLSYLRTSSLSPTHSDSRPNPPFRHHFIPHIKGNEPSSGRNSPLP--------------------------------------
Epitope (n=5)  |     ----------------PTHSDSRPNPPFRHHFIPHIKGNEPSSGRNSPLPYRPDSRPLTPTYAQA-----------------------
Epitope (n=2)  |     -------------------------PPFRHHFIPHIKGNEPSSGRNSPLPYRPDSRPLTPTYAQAPKHFHVPDQ--------------
Epitope (n=1)  |     ----------------------------------------------NSPLPYRPDSRPLTPTYAQAPKHFHVPDQGINIYRKPPIYKQH

Isoform a      | 522 YRKPPIYKQH(---)RRSSGREEDDEELLRRRQLQEEQLMKLNSGLGQLILKEEMEKESRERSS 632
Isoform j      | 246 YRKPPIYKQHGPDMKRRSSGREEDDEELLRRRQLQEEQLMKLNSGLGQLILKEEMEKESRERSS 309
Epitope (n=2)  |     --KPPIYKQHGPDMKRRSSGREEDDEELLRRRQLQEEQLMKLNSGLGQLIL-------------
Epitope (n=1)  |     --------------KRRSSGREEDDEELLRRRQLQEEQLMKLNSGLGQLILKEEMEKESRERS-

Isoform a      | 675 YNSYGDVSGGVRDYQTLPDGHMPAMRMDRGVSMPNMLEPKIFPYEMLMVTN 725
Isoform j      | 352 YNSYGDVSGGVRDYQTLPDGHMPAMRMDRGVSMPNMLEPKIFPYEMLMVTN 402
Epitope (n=1)  |     -NSYGDVSGGVRDYQTLPDGHMPAMRMDRGVSMPNMLEPKIFPYEMLMVT-
```

Figure S9. **Immunodominant peptide epitopes on ABLIM1.** Phage-displayed library sequences (epitope sequences) that were immunoprecipitated by ILD patient plasma antibodies were aligned to ABLIM1 isoform a (NP_002304) and isoform j (NP001309817). Number of patients that had reactivity to each epitope sequence is indicated on the figure. Highlighted region indicates the proline-rich region, which is commonly represented in a total of 12 phage-immunoprecipitated sequencing hits.
