## Supplemental Tables for "Antigenic responses are hallmarks of fibrotic interstitial lung diseases independent of underlying etiologies"

**Table S1.**
Flow cytometry panel for T cells

| Immunogen | Conjugate | Clone | Vendor | Dilution |
|---|---|---|---|---|
| PD1 | BV421 | MIH4 | BD | 25 |
| CD19 | BUV395 | HIB19 | BD | 100 |
| CD38 | eFluor450 | HIT2 | Invitrogen | 100 |
| CD3 | BV480 | UCHT1 | BD | 100 |
| CD45RA | BV510 | HI100 | Biolegend | 400 |
| CD8 | BV570 | RPA-T8 | Biolegend | 100 |
| Va7.2 | BV605 | 3C10 | Biolegend | 50 |
| CD127 | BV750 | HIL-7R-M2 | BD | 100 |
| HLA-DR | BV785 | L243 | Biolegend | 200 |
| CD25 | BB515 | 2A3 | BD | 200 |
| CCR7 | AF488 | G043H7 | Biolegend | 50 |
| ICOS | PerCP-Cy5.5 | C398.4A | Biolegend | 200 |
| hu CD1d PBS-57 | PE | - | NIH tetramer facility | 200 |
| CD137 | PE-Cy5 | 4B4-1 | Biolegend | 100 |
| Vd1 | PE-Cy7 | TS8.2 | Invitrogen | 50 |
| Vd2 | PE-Cy7 | B6 | Biolegend | 100 |
| CD4 | AF700 | OKT4 | Biolegend | 200 |
| BCL6 | APC-Cy7 | K112-91 | BD | 2500 |
| RORgt | BV650 | Q21-559 | BD | 50 |
| GATA3 | BV711 | L50-823 | BD | 200 |
| CD154 | APC | 24-31 | Biolegend | 50 |
| FOXP3 | AF647 | 206D | Biolegend | 50 |
| Zombie | NIR | - | BIolegend | 2500 |

**Table S2.**
Flow cytometry panel for B cells

| Immunogen | Conjugate | Clone | Vendor | Dilution |
|---|---|---|---|---|
| CD3 | PE-Cy5 | HIT3a | BD | 200 |
| HLA-DR | APC-Fire 750 | L243 | Biolegend | 333 |
| CD10 | BV711 | HI10a | Biolegend | 100 |
| CD14 | BUV395 | MΦP9 | BD | 100 |
| CD16 | BV650 | 3G8 | Biolegend | 333 |
| CD19 | PE-Cy7 | HIB19 | Biolegend | 200 |
| CD20 | eFluor450 | 2H7 | Invitrogen | 100 |
| CD24 | BV510 | ML5 | Biolegend | 200 |
| CD27 | APC-R700 | M-T271 | BD | 100 |
| CD38 | APC | HIT2 | Biolegend | 100 |
| CD44 | AF488 | BJ18 | Biolegend | 1000 |
| CD116 | BV421 | hGMCSFR-M1 | BD | 333 |
| CD138 | PE-Dazzle | MI15 | Biolegend | 50 |
| IgD | PerCP-Cy5.5 | IA6-2 | BD | 100 |
| IgM | BV785 | MHM-88 | Biolegend | 333 |
| ILT3 | AF647 | ZM4.1 | BIolegend | 333 |
| ILT5 | PE | MKT5.1 | Biolegend | 333 |
| CD71 | BUV805 | L01.1 | BD | 100 |
| CXCR4 | BUV737 | 12G5 | BD | 3333 |
| CD172 | BV750 | SE5A5 | BD | 333 |
| CD11C | BUV661 | B-ly6 | BD | 333 |
| CD1C | BV605 | F10/21A3 | BD | 333 |
| CD26 | BV480 | L272 | BD | 100 |
| Zombie | NIR | - | BIolegend | 2500 |
